# Biochemical, Biophysical, and Immunological Characterization of Respiratory Secretions in Severe SARS-CoV-2 (COVID-19) Infections

**DOI:** 10.1101/2022.03.28.22272848

**Authors:** Michael J. Kratochvil, Gernot Kaber, Sally Demirdjian, Pamela C. Cai, Elizabeth B. Burgener, Nadine Nagy, Graham L. Barlow, Medeea Popescu, Mark R. Nicolls, Michael G. Ozawa, Donald P. Regula, Ana E. Pacheco-Navarro, Samuel Yang, Vinicio A. de Jesus Perez, Harry Karmouty-Quintana, Andrew M. Peters, Bihong Zhao, Maximilian L. Buja, Pamela Y. Johnson, Robert B. Vernon, Thomas N. Wight, Stanford COVID-19 Biobank Study Group, Carlos E. Milla, Angela J. Rogers, Andrew J. Spakowitz, Sarah C. Heilshorn, Paul L. Bollyky

## Abstract

Thick, viscous respiratory secretions are a major pathogenic feature of COVID-19 disease, but the composition and physical properties of these secretions are poorly understood. We characterized the composition and rheological properties (i.e. resistance to flow) of respiratory secretions collected from intubated COVID-19 patients. We find the percent solids and protein content are greatly elevated in COVID-19 compared to heathy control samples and closely resemble levels seen in cystic fibrosis, a genetic disease known for thick, tenacious respiratory secretions. DNA and hyaluronan (HA) are major components of respiratory secretions in COVID-19 and are likewise abundant in cadaveric lung tissues from these patients. COVID-19 secretions exhibit heterogeneous rheological behaviors with thicker samples showing increased sensitivity to DNase and hyaluronidase treatment. In histologic sections from these same patients, we observe increased accumulation of HA and the hyaladherin versican but reduced tumor necrosis factor–stimulated gene-6 (TSG6) staining, consistent with the inflammatory nature of these secretions. Finally, we observed diminished type I interferon and enhanced inflammatory cytokines in these secretions. Overall, our studies indicate that increases in HA and DNA in COVID-19 respiratory secretion samples correlate with enhanced inflammatory burden and suggest that DNA and HA may be viable therapeutic targets in COVID-19 infection.

## Introduction

Severe infections of SARS-CoV-2, the virus responsible for the COVID-19 pandemic, can result in acute respiratory distress syndrome (ARDS) (1), a condition marked by viscous respiratory secretions and respiratory distress (2). The compositional and rheological properties of these respiratory secretions impair their mucociliary clearance, resulting in a build-up of fluids in the lungs during ARDS (3). This greatly inhibits oxygen exchange, often necessitating endotracheal intubation and mechanical ventilation (4). Treatments that target these respiratory secretions are desperately needed to improve clinical outcomes for COVID-19 patients as well as for other patients suffering from severe cases of ARDS. It is therefore important to understand the composition of these secretions to better guide treatment development efforts.

The typical composition of respiratory secretions consists of dilute mucins (5), which are long polymers that can form a network by entangling with each other within the aspirate fluid. Similar to mucin, other biopolymers, such as DNA and hyaluronan (HA), can form entanglements with itself and with mucin, forming more entanglements with increasing polymer concentration and contributing to a greater modulus (i.e., a greater resistance to flow). Respiratory secretions with higher modulus are expected to be more challenging to clear from the airway and hence hinder oxygen exchange in the lungs (4, 6, 7).

Levels of HA, a linear glycosaminoglycan, are elevated in respiratory secretions in other forms of respiratory inflammation (8–12) including ARDS (13–15). HA is produced at the cell surface by a variety of cell types (16) in response to viral DNA and other factors (17). HA is present in the body at molecular weights ranging from low kilodaltons to megadaltons (16, 18) and is known to have major effects on the viscoelasticity of respiratory secretions and other materials (19, 20). Additionally, HA plays important roles in innate immunity and antigenic responses in the lungs (21–24).

DNA levels are also elevated in some forms of respiratory inflammation (25, 26). This increase likely originates from dead cells, infiltrating neutrophils (27, 28), and potentially microbial contaminants (7, 29). Relatively small increases in DNA concentrations can dramatically change the rheological properties of a solution, a phenomenon that has been leveraged both naturally in the production of bacterial biofilms (30) and synthetically in the development of DNA-based hydrogels (31). In the context of lung infections, extracellular DNA has been suggested to increase viscosity of mucosal fluid and provide colonization opportunities for bacterial infections (25).

We hypothesized that DNA and HA are major contributors to the tenacious behavior of respiratory secretions from COVID-19 patients. To evaluate this, we characterized the composition, rheological properties (e.g., viscosity and elasticity properties), and cytokine/chemokine profiles of respiratory secretions from patients under mechanical ventilation due to severe COVID-19, given the importance of these parameters in other respiratory diseases (32, 33). As controls for these studies, we have included sputum samples from both healthy individuals and subjects with cystic fibrosis (34), a genetic disease associated with notoriously thick lung secretions (35). These studies provide biochemical, biophysical, and immunological assessment of respiratory secretions in COVID-19.

## Results

### Solids and proteins are increased in COVID-19 respiratory secretions

We collected respiratory secretions from ventilated COVID-19 patients, ranging from 5 to 70 years of age (Table 1). These were collected via suction catheter, with only a single sample from each individual included in the dataset. Respiratory secretion samples were collected from patients with CF via spontaneous expectoration and from healthy volunteers via sputum induction.

**Table 1.** Clinical Characteristics of SARS-CoV2+ patients.

| Clinical Characteristic | Adult Patients<br>(n=22) | Pediatric Patients<br>(n = 2) |
| --- | --- | --- |
| Age (years) | 57 (41 - 69) | 5-15 |
| Male gender | 15 (68%) | 2 (100%) |
| Race/Ethnicity | Non-black Hispanic 13 (59%)<br>White 3 (14%)<br>Black 2 (9%)<br>Asian 3 (14%)<br>Pacific Islander 1 (5%) | Non-black<br>Hispanic 1 (50%)<br>Pacific Islander 1<br>(50%) |
| Preexisting diabetes<br>mellitus | 12 (55%) | 0 |
| Preexisting pulmonary<br>disease | 4 (18%) | 0 |
| Preexisting cardiac disease | 4 (18%) | 1 (50%) |
| Hospital length of stay | 34.5 (22-47) | 113, 291+* |
| In-hospital death | 8 (36%) | 0 |
| Total ventilator days | 19.6 (9-36) | 9, 290+* |
| In-hospital tracheostomy | 7 (32%) | 1 (50%) |
| Days from intubation to<br>first collection | 2 (2-4.6) | 1, 16 |
| ARDS Severity at time of<br>collection | Mild 1 (7%)<br>Moderate 14 (64%)<br>Severe 5 (23%)<br>Recovered 2 (14%) | Moderate 1 (50%)<br>Severe 1 (50%) |
| Antibiotics at time of<br>collection | 12 (55%) | 0 |

For adult patients, values listed as N (%) or Median (IQR). In pediatric patients, values listed as N (%) or given for both patients. \*Patient still admitted and mechanically ventilated at time of publication.

We observed that samples from healthy patients were typically clear and colorless, whereas samples from patients with COVID-19 were typically colored and opaque, similar to samples from patients with CF (Figure 1A), This suggested that the samples contain appreciable amounts of biopolymers and non-soluble debris.

**Figure 1.**
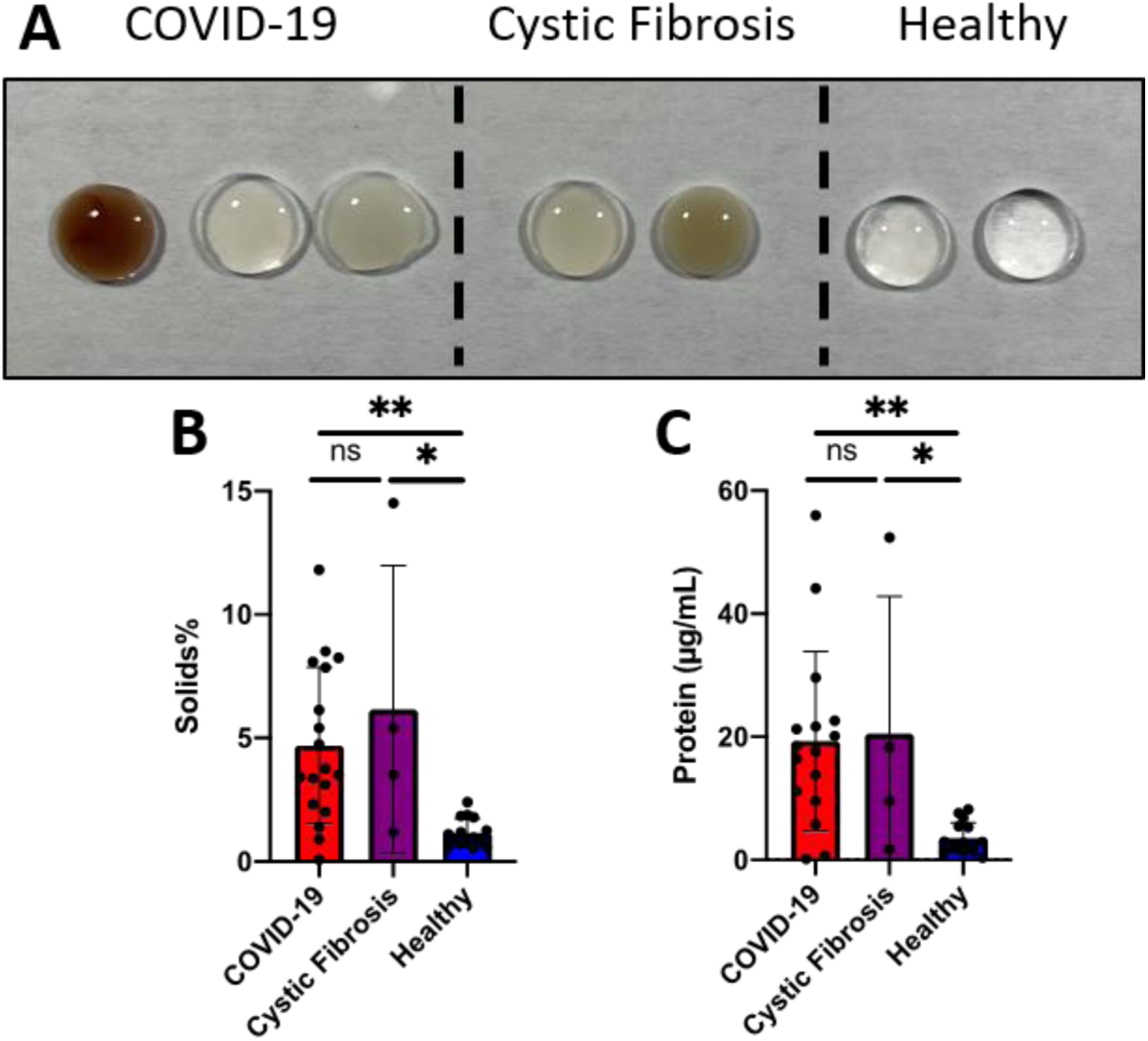
Respiratory secretions from patients with COVID-19 are high in solids and protein compared to healthy subjects. (**A**) Representative images of respiratory secretions collected from ventilated patients with COVID-19 are viscous and tenacious. Similar to CF samples, COVID-19 samples are often colored and opaque, whereas healthy samples are clear and colorless. (**B**) Quantification of solids found in COVID-19 (n=18), CF (n=4), and healthy (n=15) respiratory secretions. (**C**) Quantification of protein concentration in COVID-19 (n=16), Cystic Fibrosis (n=4), and healthy (n=15) respiratory secretions. One-way ANOVA with Tukey multiple comparisons tests. *p<0.05, **p<0.01.

The percent solids content of respiratory secretions, an index of hydration, impacts the difficulty with which respiratory secretions can be cleared and correlates with clinical outcomes in CF and other settings (36–38). We found that COVID-19 samples had significantly higher percent solids than healthy samples (Figure 1B). We further observed that protein concentrations in COVID-19 samples were nearly 5.5 times greater than those seen in healthy samples (Figure 1C, p = 0.003). COVID-19 and CF samples did not show statistically significant differences (p = 0.983). These data are consistent with infected and inflamed lungs being known to have protein deposits from increased mucin production (39), bacterial colonization (40), and infiltrating cells (28).

Of note, large variances in solids and protein were observed in the COVID-19 patient samples (Figure 1B-C). Even though all the patient samples were collected from intubated patients with severe COVID-19 early during mechanical ventilation, this variance may reflect the differences in the individual response to the infection and the disease progression at the time of collection.

### HA is increased in COVID-19 respiratory secretions and lung sections

We next evaluated HA content in the respiratory secretions. We observed a statistically significant, 10-fold increase in HA concentration in COVID-19 patient samples compared to samples from healthy subjects (Figure 2A, p = <0.0001). The average concentration of HA found in samples from COVID-19 subjects was comparable to that observed in samples from CF subjects (p = 0.333), a disease state associated with greatly increased sputum HA (41). Similar to our findings with percent solids and protein, we observed larger variance in the amounts of HA in COVID-19 and CF patient samples than compared to samples from healthy donors.

**Figure 2.**
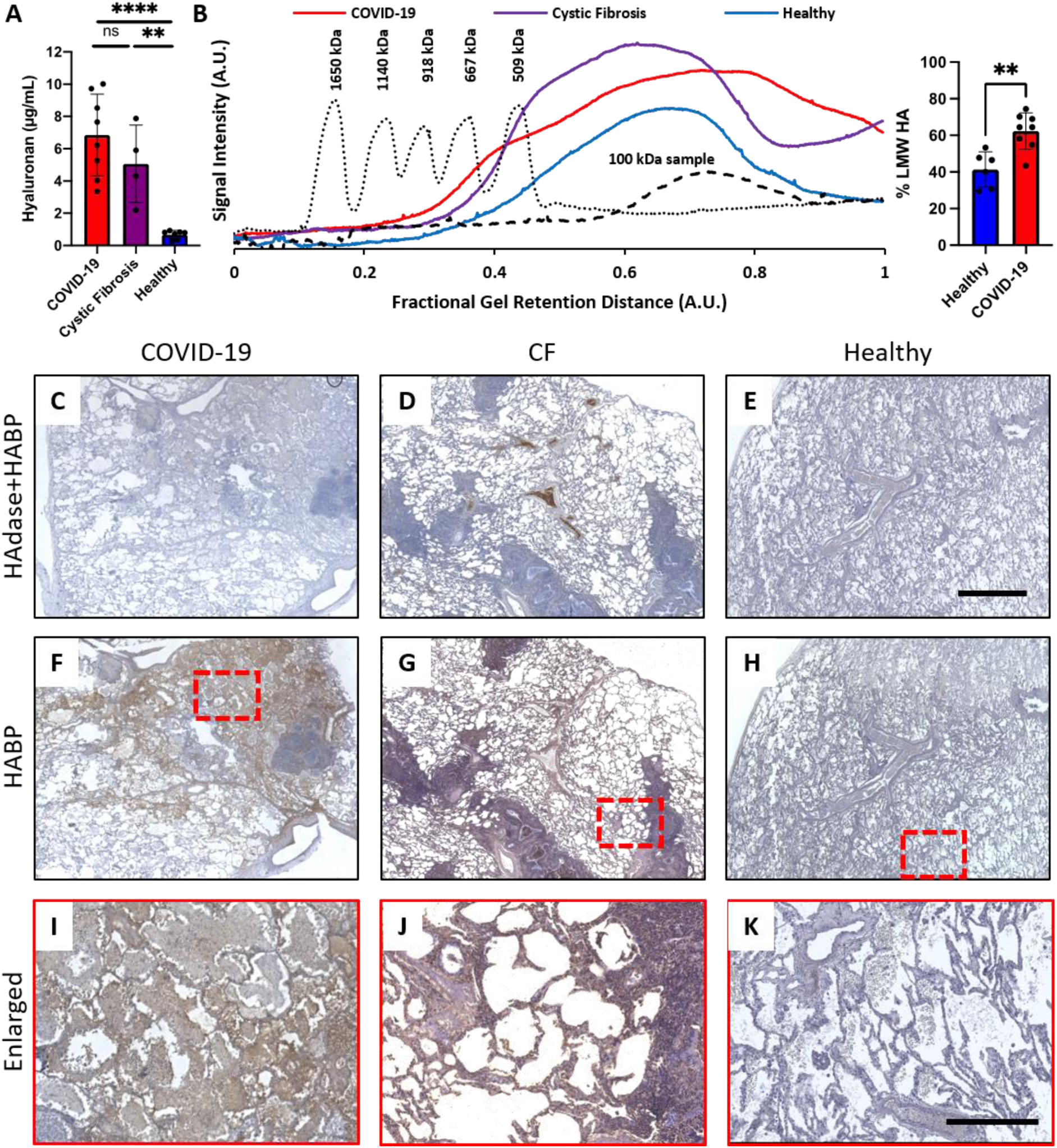
COVID-19 human lung sections have high levels of HA. (**A**) Quantification of HA in respiratory secretion samples. COVID-19 (n=8), CF (n=4), Healthy (n=7) respiratory secretion samples. One-way ANOVA with Tukey multiple comparisons tests; **p<0.01, ****p<0.0001. (**B**) Representative chromatogram of HA molecular weight (MW). Solid traces are the averages of COVID-19, CF, and healthy respiratory secretion samples. The dotted trace is the chromatogram of standard loaded with HA of known MWs, as indicated on graph. The dashed trace is representative of a commercially available 100 kDa MW HA. The bar graph represents the % low MW HA in respiratory secretion samples (Healthy (n=6), COVID-19 (n=8)). Unpaired t test with Welch’s correction; **p<0.005. (**C-H**) Representative histological cadaveric lung sections from donors with COVID-19, donors with CF, and healthy donors, both with (**C-E**) and without (**F-H**) HAdase treatment. Nuclei are stained in blue, and HA binding proteins (HABP) are stained in brown. (**I-K**) Enlarged sections from panels F-H, respectively. Scale bars C-H 800 µm, I-K 400 µm.

Given that the molecular weight of HA is known to influence both the immunogenic as well as the rheological properties of the resulting solution (42–44), we measured the molecular weight of the HA in the different samples (Figure 2B; Supplemental Figure 1). We found that while all samples of respiratory secretions had HA of molecular weight less than 500 kDa, HA size in samples from patients with COVID-19 donors skewed smaller than that seen in samples from donors with CF and healthy controls. Given that low molecular weight HA polymers promote inflammation in some systems (42, 43) this is consistent with the highly inflammatory nature of respiratory disease in COVID-19 infection.

### HA is increased in COVID-19 cadaveric lung sections

We next examined cadaveric lung sections from patients with COVID-19, patients with CF, and patients with healthy lungs (i.e., without a diagnosed pulmonary disease) for HA deposits by staining with HA binding protein (HABP). We observed very little HABP staining in sections treated with hyaluronidase (HAdase) (Figure 2C-E), suggesting very low non-specific background staining. However, we observed a substantial increase in HA staining in lung sections from both COVID-19 and CF donors compared to healthy samples when no prior HAdase treatment was used (Figure 2F-H). We observed strong diffuse staining in areas of necrosis and inflammation. Higher magnification images demonstrated the accumulation of HA within alveolar spaces (Figure 2I-K). These data, together with the aforementioned respiratory secretion studies, indicated that patients with severe COVID-19 have elevated levels of HA in their lungs.

### HA and hyaladherins are increased in blood vessels of COVID-19 lung sections

We next determined whether changes in HA accumulation corresponded to changes in hyaladherins, the extracellular matrix molecules that bind and interact with HA, including versican and tumor necrosis factor–stimulated gene-6 (TSG-6). The donor profiles for these histologic tissue samples are included in Table 2.

**Table 2.** Demographic details for histology studies.

| **Sample ID** | **Age** | **Sex** | **Group** |
| --- | --- | --- | --- |
| **1** | 30's | F | Healthy |
| **2** | 60's | M | Healthy |
| **3** | 60's | M | Healthy |
| **4** | 50's | M | Healthy |
| **5** | 50's | M | Healthy |
| **6** | 50's | F | COVID-19 ARDS |
| **7** | 50's | F | COVID-19 ARDS |
| **8** | 50's | M | COVID-19 ARDS |
| **9** | 60's | M | COVID-19 ARDS |
| **10** | 70's | M | COVID-19 ARDS |
| **11** | 20's | M | Non-COVID-19 ARDS |
| **12** | 20's | M | Non-COVID-19 ARDS |
| **13** | 40's | F | Non-COVID-19 ARDS |
| **14** | 30's | M | Non-COVID-19 ARDS |
| **15** | 30's | F | CF |
| **16** | 15-20 | F | CF |
| **17** | 70's | F | CF |

The expression and accumulation of versican and TSG-6 increase significantly during inflammation in many diseases (45). We evaluated HABP, versican, and TSG-6 staining patterns (localization and intensity) in lung sections from patients with COVID-19 ARDS, non-COVID-19 ARDS, CF, and patients with healthy lungs (patients who died without known lung disease). Compared to healthy control samples, both COVID-19 ARDS and non-COVID-19 ARDS lung sections demonstrate intense HABP staining of the distorted alveolar-capillary barrier, including HABP staining in the alveolar spaces (Figure 3A-C) (Supplemental Figure 2). We observed diffuse alveolar hemorrhage, distended alveolar-capillary membrane with strong heterogeneous staining, and diffuse bronchial and vascular staining in the COVID-19 lung sections. We did not observe major differences in HABP staining between these two patient groups. In comparison, for the CF samples, HABP staining was mostly seen in inflammatory cells, mucus, and the medial layer of bronchi and vessels. Far more HA staining was observed in all of these conditions than in tissues from subjects who died without lung disease (Figure 3D, M) (Supplemental Figure 2).

**Figure 3.**
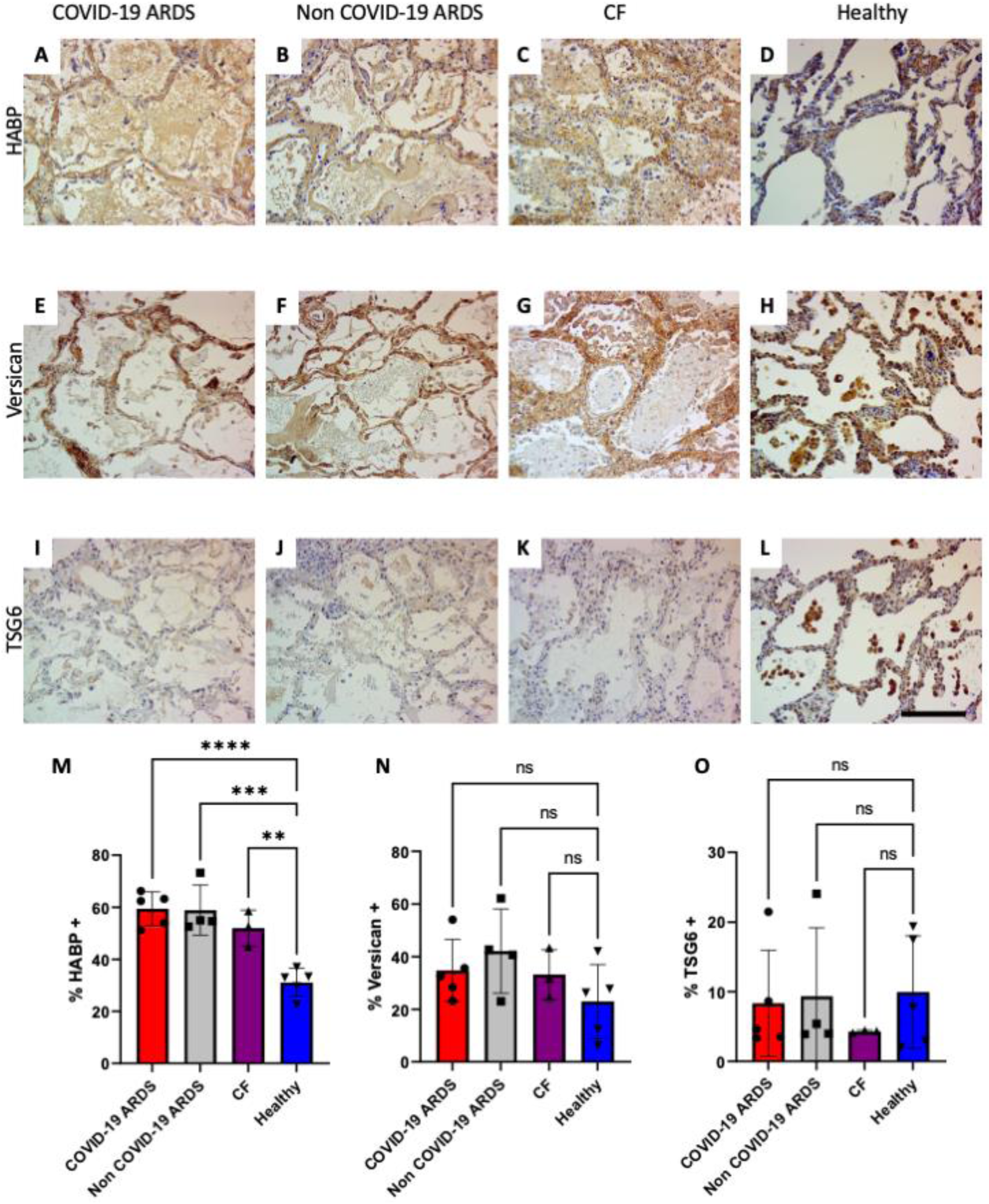
HA and hyaladherins are increased in blood vessels of COVID-19 lung sections. Representative histological cadaveric lung sections from donors with COVID-19 ARDS, donors with non-COVID-19 ARDS, donors with CF, and healthy donors stained with (**A-D**) HABP, (**E-H**) versican, (**I-L**) TSG6 (40X magnification). Nuclei are stained in blue, and HABP, versican, or TSG6 are stained in brown. Scale bar A-L 400 µm. Tissues were examined using an Amscope T720Q microscope and images (40X) were acquired using Amscope digital camera (MU1403) and imaging software. (**M**) % HABP+, (**N**) % versican+, and (**O**) % TSG6+ area in lung sections from COVID-19 ARDS (n=5), non-COVID-19 ARDS (n=4), CF (n=3), and healthy donors (n=5). One-way ANOVA with Dunnett’s multiple comparisons tests; **p<0.01, **p<0.001, ****p<0.0001.

In healthy lung tissues, versican staining is strong in alveolar macrophages, bronchial epithelia, and peribronchial layer, with some staining in the medial layer of blood vessels. In contrast, both COVID-19 ARDS and non-COVID-19 ARDS samples display much stronger and prominent versican staining in the medial layer of blood vessels and the alveolar-capillary membrane, compared to healthy controls. Similar findings occur in CF except that strong staining is also seen in the bronchial epithelium (Figure 3E-H) (Supplemental Figure 3). Although the total accumulation of versican is not significantly different (trending higher) in COVID-19 ARDS, non-COVID-19 ARDS, and CF lung sections, compared to healthy controls (Figure 3N), the tissue distribution and localization is altered as described above and in Table 3.

**Table 3.** Histologic staining results of lung tissues for HA, versican, and TSG6.

| **Patient identifier** | **Stain** | **Group** | **Histopathology results** |
| --- | --- | --- | --- |
| **1** | HABP-B | Healthy | Mild staining of the alveolar-capillary membrane. Strong adventitia and peribronchial staining. |
| **1** | Versican | Healthy | Strong bronchial epithelial staining, mild to moderate peribronchial staining, mild perivascular stain |
| **1** | TSG6 | Healthy | Strong staining of bronchial epithelium and alveolar macrophages. Some alveolar epithelial cells also show mild staining. |
| **2** | HABP-B | Healthy | Moderate staining in the bronchial wall, vascular adventitia and alveolar-capillary membrane. Alveolar macrophages also exhibit strong intracellular staining. |
| **2** | Versican | Healthy | Strong adventitial stain around medium around small pulmonary arteries. No bronchial epithelium stain. Medium to strong peribronchial staining. |
| **2** | TSG6 | Healthy | Strong staining of bronchial epithelium and alveolar macrophages. Mild to medium stain of medial layer of pulmonary arteries. Strong alveolar epithelial staining. |
| **3** | HABP-B | Healthy | Strong staining in the bronchial wall, vascular adventitia and alveolar-capillary membrane. Mild to moderate staining in the basement membrane of medium size vessels. |
| **3** | Versican | Healthy | Strong alveolar macrophage staining. Mild stain of pulmonary veins. Strong bronchial epithelial and peribronchial staining. Strong staining in basement membrane and adventitia of pulmonary arteries. |
| **3** | TSG6 | Healthy | Strong bronchial epithelial stain, Strong alveolar macrophage stain, mild to moderate alveolar-capillary stain, no stain of the small pulmonary vessels (50um); mild subendothelial staining of medium size vessels. |
| **4** | HABP-B | Healthy | Strong staining in the bronchial wall, veins, vascular adventitia and alveolarcapillary membrane. Mild to moderate staining in the basement membrane of medium size vessels. |
| **4** | Versican | Healthy | Strong alveolar macrophage staining. Mild stain of pulmonary veins. Strong bronchial epithelial and peribronchial staining. Strong staining in basement membrane and adventitia of pulmonary arteries. |
| **4** | TSG6 | Healthy | Strong staining of bronchial epithelium and alveolar macrophages. Some alveolar epithelial cells also show mild staining. |
| **5** | HABP-B | Healthy | Relative well preserved lung tissue with minimal peribronchial inflammation. Staining can be seen in parenchyma with stronger staining in airway and vascular media. |
| 5 | Versican | Healthy | Faint stain throughout, stronger in internal and external elastic lamina of blood vessels. |
| 5 | TSG6 | Healthy | Faint stain, relatively well preserved parenchyma, some staining in the intimal layer of blood vessels, strong stain in perivascular inflammatory cells |
| 6 | HABP-B | COVID-19 ARDS | Lung shows evidence of extensive consolidation with epithelial detachment and hemorrhage. Strong diffuse staining in areas of necrosis and inflammation. Strong staining across the entire wall of medium sized vessels. |
| 6 | Versican | COVID-19 ARDS | Strong stain adjacent to the external elastic lamina of small and medium size arteries and veins, strong stain in the basement membrane of alveolar epithelial cells. |
| 6 | TSG6 | COVID-19 ARDS | Mild to moderate staining of the alveolar macrophages and other immune cells infiltrating the parenchyma. Moderate to strong staining of alveolar epithelium and endothelial layer of vessels. |
| 7 | HABP-B | COVID-19 ARDS | Lung shows complete consolidation with epithelial detachment, hemorrhage, and necrosis. No intact alveolar structure can be appreciated. Strong staining across the entire wall of medium sized vessels. |
| 7 | Versican | COVID-19 ARDS | Strong stain adjacent to the external elastic lamina of small and medium size arteries and veins, Large vessels show strong staining across the entire wall. |
| 7 | TSG6 | COVID-19 ARDS | Moderate to strong staining of immune cells infiltrating the parenchyma. Moderate to strong staining of endothelial layer of vessels. |
| 8 | HABP-B | COVID-19 ARDS | Diffuse alveolar hemorrhage, distended alveolar-capillary membrane with strong heterogeneous staining; diffuse bronchial and vascular staining. |
| 8 | Versican | COVID-19 ARDS | Strong stain within medial layer of large to small size arteries, moderate to strong staining in the alveolar-capillary membrane. |
| 8 | TSG6 | COVID-19 ARDS | Moderate staining of immune cells infiltrating the parenchyma. Mild staining of medial layer of vessels. Staining in alveolar-capillary membrane likely represents immune cells. |
| 9 | HABP-B | COVID-19 ARDS | Diffuse alveolar hemorrhage, distended alveolar-capillary membrane with strong heterogeneous staining; diffuse bronchial and vascular staining. |
| 9 | Versican | COVID-19 ARDS | Strong stain within medial layer of large to small size arteries, mild staining in the alveolar-capillary membrane. One large vessels with diffuse strong staining can be seen at the center of the slide. |
| 9 | TSG6 | COVID-19 ARDS | Mild stain of the alveolar macrophages and other immune cells. |
| **10** | HABP-B | COVID-19 ARDS | Diffuse alveolar hemorrhage, distended alveolar-capillary membrane with strong heterogeneous staining; diffuse bronchial and vascular staining. |
| **10** | Versican | COVID-19 ARDS | Strong stain within the subendothelial and adventitial layer of large to small size arteries, moderate to strong staining in the alveolar-capillary membrane. |
| **10** | TSG6 | COVID-19 ARDS | Mild stain of the alveolar macrophages and other immune cells. Heterogeneous stain of intima and adventitia of medium sized vessels. |
| **11** | HABP-B | non-COVID-19 ARDS | Strong signal in medium pulmonary arteries, particularly in the adventitial layer, and immune cells. Mild to moderate stain in the alveolar-capillary membrane |
| **11** | Versican | non-COVID-19 ARDS | Strong stain within medial layer of large to small size arteries, moderate staining in the alveolar-capillary membrane. |
| **11** | TSG6 | non-COVID-19 ARDS | Mild staining of alveolar macrophages and other immune cells. |
| **12** | HABP-B | non-COVID-19 ARDS | Diffuse stain across the lung tissue. No discernible structures in most of the sample due to massive cell immune infiltration and tissue destruction. |
| **12** | Versican | non-COVID-19 ARDS | Strong stain within medial layer of large to small size arteries, mild staining in the alveolar-capillary membrane. |
| **12** | TSG6 | non-COVID-19 ARDS | Mild staining of alveolar macrophages and other immune cells, Scattered staining of endothelial cells in medium size vessels. |
| **13** | HABP-B | non-COVID-19 ARDS | Strong signal intensity in the alveolar-capillary membrane. Alveolar macrophages and other immune cells within alveoli have mild or no stain. |
| **13** | Versican | non-COVID-19 ARDS | Strong stain within medial layer of large to small size arteries, mild to moderate staining in the alveolar-capillary membrane. |
| **13** | TSG6 | non-COVID-19 ARDS | Strong staining of alveolar macrophages and other immune cells. Strong staining of alveolar epithelium. |
| **14** | HABP-B | non-COVID-19 ARDS | Strong signal intensity in the alveolar-capillary membrane. Strong perivascular staining. |
| **14** | Versican | non-COVID-19 ARDS | Strong stain within medial layer of large to small size arteries, moderate to high staining in the alveolar-capillary membrane. |
| **14** | TSG6 | non-COVID-19 ARDS | Mild staining of alveolar macrophages and other immune cells. |
| **15** | HABP-B | CF | Diffuse stain -stronger around medial layer of blood vessels and airways |
| **15** | Versican | CF | Strong stain of the medial layer of medium and small muscularized vessels. Staining can also be seen inside alveolar type 1 and 2 epithelial cells |
| **15** | TSG6 | CF | Faint staining throughout; strong stain in subendothelial layer, strongest in inflammatory cells within alveoli. |
| **16** | HABP-B | CF | Diffuse stain -stronger around medial layer of blood vessels and airways; several inflammatory clusters. |
| **16** | Versican | CF | Strong stain of the medial layer of medium and small muscularized vessels; strong stain in bronchial submucosa. |
| **16** | TSG6 | CF | Strong stain on mucus and bronchial epithelial cells. |
| **17** | HABP-B | CF | Diffuse stain -stronger around medial layer of blood vessels and airways, mucus pools seen in alveoli and bronchial lumen also stain positive. |
| **17** | Versican | CF | Diffuse parenchymal staining, strong stain in bronchial mucosa around engorged glands. |
| **17** | TSG6 | CF | Strong stain in bronchial epithelium, inflammatory cells, medium stain in vascular media. |
| **18** | TSG6 | Lung tumor | Staining controls |
| **18** | Rbt IgG | Lung tumor | Staining controls |
| **19** | TSG6 | Kidney (Pan QC0020A3) | Staining controls |
| **20** | Versican | Human lung tumor | Staining controls |
| **20** | Rbt IgG | Human lung tumor | Staining controls |
| **21** | Versican | DIVA CF lung | Staining controls |
| **21** | Rbt IgG | DIVA CF lung | Staining controls |

For TSG6, we observed strong staining of bronchial cells and immune cells in lung sections from healthy, CF, and non-COVID-19 ARDS. However, interestingly, in COVID-19 ARDS samples, the stain is much fainter in alveolar macrophages and other immune cells (Figure 3I-L) (Supplemental Figure 4). We also quantified the total TSG6+ staining in all of the lung sections and observed no significant change in the overall accumulation of TSG6 (Figure 3O). The full review of each stained histology slide is included in Table 3.

Overall, for both COVID-19 ARDS and non-COVID-19 ARDS, there is a shift towards stronger staining of versican in the blood vessels, which correlates with the severity of tissue damage. There also seems to be more evidence of alveolar hemorrhage associated with strong and diffuse HA staining in alveolar spaces of COVID-19 ARDS lung sections.

### DNA is increased in COVID-19 respiratory secretions

We next examined the double-stranded DNA (dsDNA) content in these respiratory secretion samples. The respiratory secretions collected from patients with COVID-19 had increased dsDNA content compared to healthy subjects (Figure 4A, p = 0.032). The average dsDNA content in the COVID-19 samples was 14 times greater than that in the healthy samples, with eight of the observed samples having over ten times more dsDNA. CF sputum, by comparison, had roughly a comparable average dsDNA concentration to COVID-19 samples (p = 0.999). Sizing the dsDNA in the samples suggested that the dsDNA is very large (greater than 10 kb, i.e. >6,000 kDa) (Figure 4B; Supplemental Figure 5). The variance observed in samples from patients with COVID-19 and CF was again very large in comparison to that observed in healthy donor samples.

**Figure 4.**
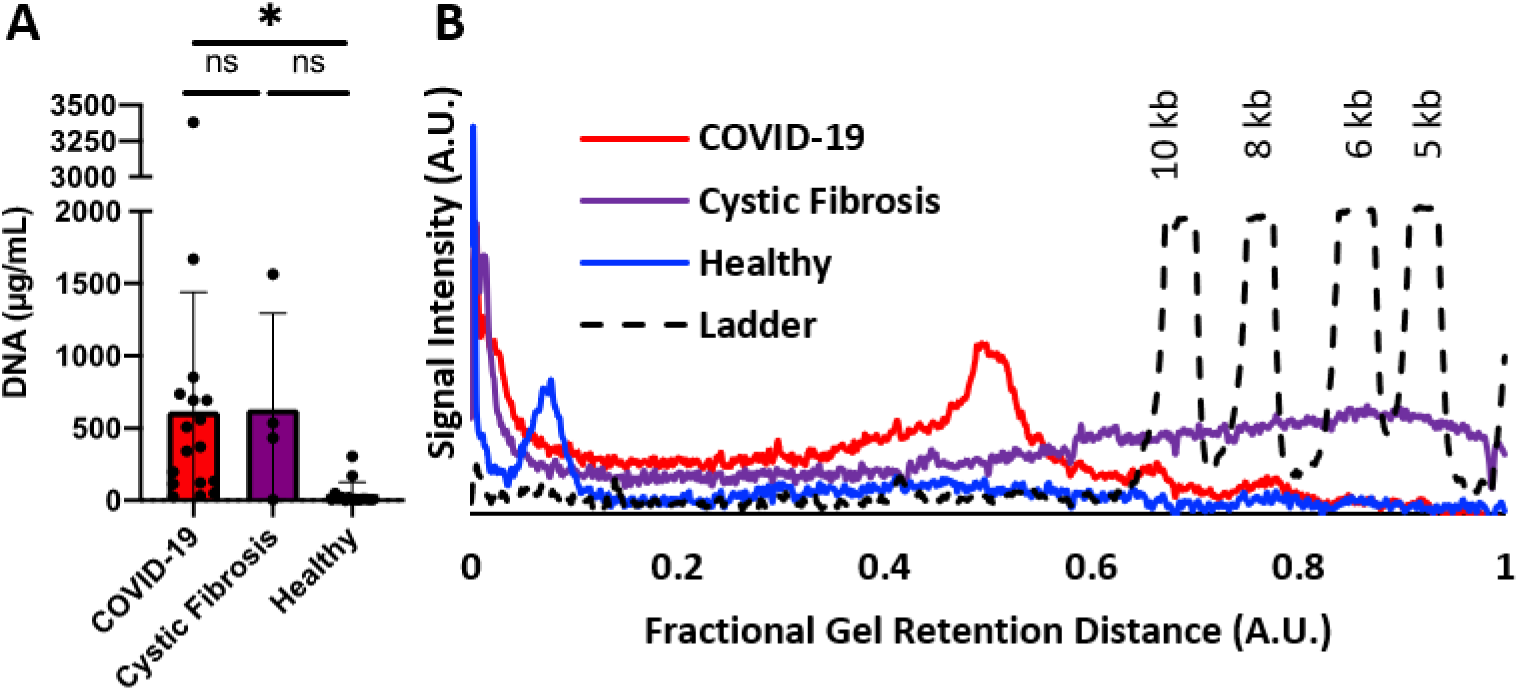
Increased levels of dsDNA in COVID-19 respiratory secretions. (**A**) Quantification of dsDNA in respiratory secretion samples. COVID-19 (n=17), CF (n=4), Healthy (n=15) respiratory secretion samples. One-way ANOVA with Tukey multiple comparisons tests. *p<0.05. (**B**) Representative chromatogram of dsDNA molecular weight. Solid traces are the averages of COVID-19, CF, and healthy respiratory aspirate samples. The dashed line trace is the chromatogram of a DNA standard ladder with the dsDNA base pair-lengths labeled above the respective peaks.

### High modulus secretions are more susceptible to enzymatic treatment

We next evaluated the rheological properties of COVID-19 respiratory secretions and the contribution of HA and dsDNA to the physical flow properties of the secretions. These flow properties can be expected to greatly impact the ability of patients of to clear secretions from the lungs. Dynamic light scattering microrheology, a non-invasive rheology technique, was used to evaluate the rheological properties of the sample due to the small sample volume required and the ability of the technique to not alter the sample properties during measurement (46, 47). The samples were measured both before and following enzymatic treatment (microrheology protocol further described in Methods and Supplemental Figures 6-7). Specifically, we examined the impact of enzymatic treatment with HAdase (to degrade HA) or deoxyribonuclease (48) (DNase; to degrade dsDNA) on the flow properties of respiratory secretions. We hypothesized that enzymatic degradation of these biopolymers would lower the modulus (i.e. the resistance to flow) given the abundance of DNA and HA in these samples. As a non-enzymatic treatment control, samples were diluted with an equivalent volume of saline.

We evaluated the absolute impact of enzymatic treatment as a function of the measured pre-treatment modulus of the respiratory secretions (Figure 5). Samples that had a higher pre-treatment modulus (i.e. thicker samples that were more resistant to flow) had a larger response to enzymatic treatment by either DNase or HAdase compared to a control saline dilution. We found a statistically significant linear relationship between the pre-treatment modulus of the secretions and the difference between the change of modulus with dilution and change of modulus with enzymatic treatment (ΔG_Saline_ – ΔG_Enzyme_). If the enzyme had no effect compared to the dilution control, then ΔG_Saline_ – ΔG_Enzyme_ = 0; if the enzyme treatment decreased the modulus of the sample, then ΔG_Saline_ – ΔG_Enzyme_ < 0. The Bayesian Information Criterion for comparing this linear model against a fit for random noise was much greater than 10 (BIC = 35.75 for DNase, and BIC = 36.92 for HAdase), indicating strong statistical significance for this linear relationship. These data are consistent with the hypothesis that thicker COVID respiratory secretions are more sensitive to enzymatic treatments that degrade DNA and HA, resulting in a lower modulus (i.e. less resistance to flow). By comparison, healthy control samples had low pre-treatment moduli and were not dramatically impacted by enzymatic treatments.

**Figure 5.**
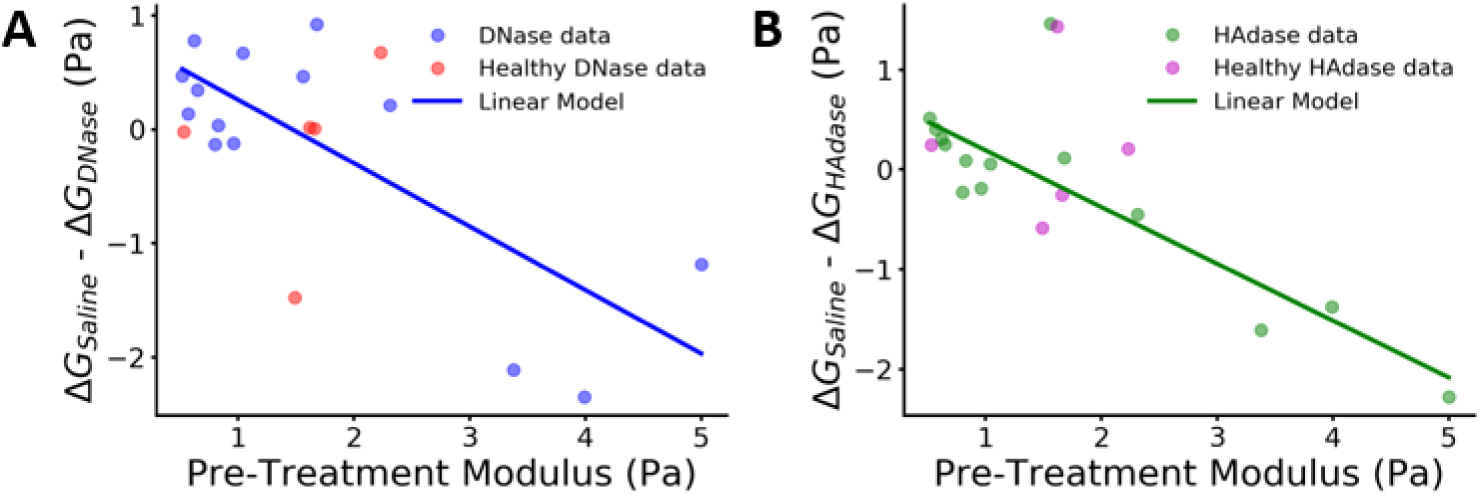
Enzymatic treatments impact the rheology of COVID-19 respiratory secretions in proportion to their pre-treatment moduli. (**A**) The difference in modulus of COVID-19 lung secretions (n=15) upon control saline dilution and enzymatic DNase treatment (ΔG_Saline_-ΔG_DNase_) versus the initial, pre-treatment modulus (blue). Healthy controls shown for comparison (n=6) (81). (**B**) The difference in modulus of COVID-19 lung secretions (n=15) upon control saline dilution and enzymatic HAdase treatment (ΔG_Saline_-ΔG_HAdase_) versus the initial, pre-treatment modulus (green). Healthy controls shown for comparison (n=6) (magenta). The Bayesian Information Criterion (BIC) was used as the statistical metric to compare the impact of enzymatic treatment. (BIC = 35.75 for DNase, and BIC = 36.92 for HAdase).

We then evaluated the relationship between the pre-treatment concentration of DNA and HA on the modulus change due to enzymatic treatment. Interestingly, we did not observe a correlation between biopolymer concentration and modulus change, as indicated by the BIC values being less than 10 (3.5 and 8.5 for DNA and HA, respectively (Supplemental Figure 8). This suggests that the contribution of HA and DNA to the modulus of these samples is complex and may be influenced by the presence of HA-binding proteins in these samples, for example.

In addition to considering the absolute reduction in the modulus as a function of the pre-treatment modulus, we also examined the percent change from the initial pre-treatment modulus. When compared to dilution, there was a trend towards a lower average modulus following enzymatic treatment with either HAdase or DNase but this did not reach statistically significance (Supplemental Figure 4). However, this approach does not account for the large variance in the modulus of pre-treatment samples, a factor which is considered in the analysis shown in Figure 5.

### Mucin expression is heterogeneous in COVID-19 respiratory secretions

We also assessed mucin glycoprotein content in the COVID-19 respiratory secretion samples. Mucins are high molecular weight (HMW) and heavily glycosylated proteins lining mucosal surfaces that play an important role in innate defense, protecting the epithelium against invading pathogens (49). The major secreted airway mucins or gel-forming mucins are Mucin 5AC (MUC5AC) and Mucin 5B (MUC5B), which are produced by goblet cells and mucous cells within submucosal glands, respectively (50). We show representative results of MUC5AC and MUC5B expression following agarose gel electrophoresis in respiratory secretion samples from healthy, COVID-19, and CF patients (Supplemental Figure 9). We observed highly variable MUC5AC and MUC5B expression in COVID-19 group, with samples ranging from undetectable expression, low expression, or high expression, compared to the healthy controls. Of note, in CF, mucin is reported to decrease during stable disease but increase during a pulmonary exacerbation (51, 52), which is consistent with our results.

One potential explanation for our COVID-19 results is that after acute infection, there may be less mucus production in some subjects due to damage of mucus producing glandular epithelial cells (49). In contrast to our study, in one published report, higher levels of MUC5AC (as measured by ELISA) were observed in airway mucus aspirated by bronchoscopy from COVID-19 ICU patients compared to induced sputum from healthy controls(53). Additional studies reported accumulation of mucins in COVID-19 patients, without comparison to mucins from healthy individuals (54, 55).

Overall, in our studies, we observed heterogeneous mucin expression in respiratory secretions from COVID-19 ARDS patients, compared to healthy controls.

### Enhanced inflammatory burden in COVID-19 respiratory secretion samples

While substantial literature exists concerning systemic immune responses and inflammatory profile during SARS-CoV-2 infection derived from serum or plasma samples, there is very little information about the inflammatory profile of respiratory secretions in COVID-19. To better understand the lung tissue specific immune responses, we measured 80 cytokines, chemokines, adhesion molecules, and growth factor levels in COVID-19 respiratory secretion samples, compared to healthy controls.

We observed significant decreases in antiviral type I interferon IFN-α2 and PDGFAA in COVID-19 respiratory secretion samples, compared to the healthy controls (Figure 6; Supplemental Figure 10). In contrast to a previous report, IL-13 was not significantly different from healthy controls in our samples (56). We observed significant upregulation of several other cytokines and chemokines in the COVID-19 cohort compared to healthy controls, including IL6, tumor necrosis factor (TNF), IFNy, IL10, IL1β, IL18, MCSF, RANTES/CCL5, MIP-1β/CCL4, MIP-1 α/CCL3, and others. Although there is heterogeneity in the response among samples within a group, groups of samples (healthy or COVID-19) follow similar trends. These findings extend previous reports describing diminished type I interferon and hyperinflammatory responses in the context of severe COVID-19 disease but show that lung cytokine responses are regulated in a distinct fashion. In contrast, we find that CF patients have more generalized elevations in their sputum cytokine and chemokine profiles (Supplemental Figure 11).

**Figure 6.**
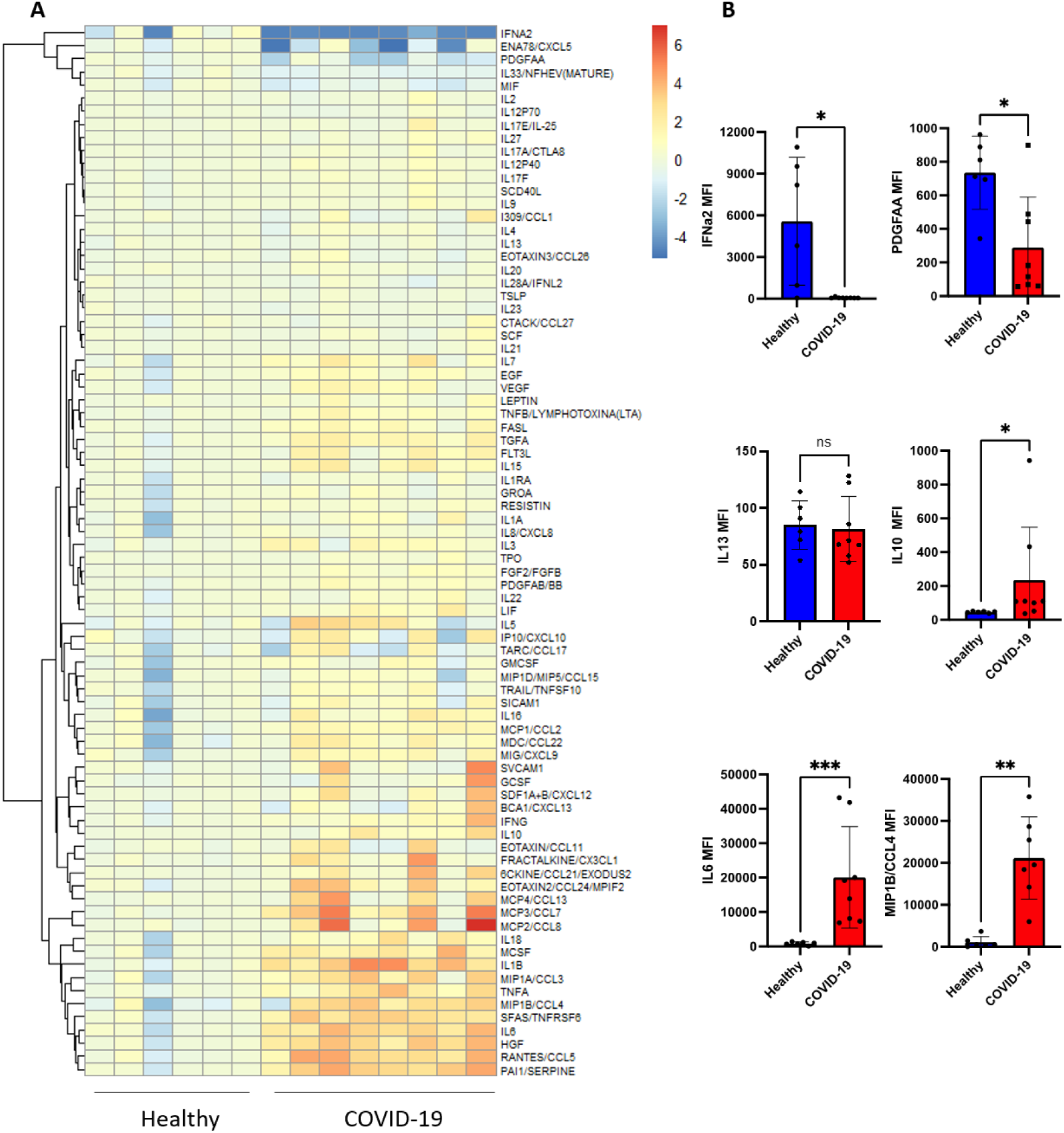
Immunological characterization of respiratory secretions from COVID-19 ARDS patients. (**A**) Heat map of mean fluorescence intensity (MFI) data, log2 transformed, and normalized to average of the healthy controls per cytokine. Data are ordered by KNN clustering of the cytokines (y-axis). Cytokines, chemokines, adhesion molecules, and growth factors were measured in the respiratory secretion samples of healthy controls (n = 6) and in COVID-19 ARDS patients (n=8) using a bead-based multiplexed immunoassay system, Luminex-EMD Millipore Human 80 Plex assays. Upregulated cytokines are shown in orange and downregulated in blue. (**B**) Bar graphs of raw MFI values for representative cytokines (IFNa2, PDGFAA, IL13, IL10, IL6, and MIP1B/CCL4) in healthy control and COVID-19 ARDS group (not normalized). Mann Whitney test; *p<0.05, **p<0.005, ****p<0.0005.

Overall, our data support that increases in HA and DNA in COVID-19 ARDS respiratory secretion samples correlate with enhanced inflammatory burden in COVID-19.

## Discussion

We report that respiratory secretions from patients with COVID-19 ARDS are thick and tenacious, comparable to the notoriously thick and tenacious sputum produced by patients with CF. COVID-19 respiratory secretions have significantly elevated levels of solids, with HA and DNA contributing to the elevated viscosity. Low-molecular weight HA in particular is greatly increased in the respiratory secretion samples from intubated patients with COVID-19. Consistent with these findings in respiratory secretions, HA is abundant in histologic sections from cadaveric lung tissues from individuals with COVID-19-associated ARDS. Together these data indicate that low-molecular weight HA is elevated in the respiratory secretions of patients with COVID-19-associated ARDS.

We found that samples that had a higher pre-treatment modulus (i.e. thicker samples that were more resistant to flow) had a larger response to enzymatic treatment by either DNase or HAdase compared to a control saline dilution. However, this change did not scale with the pre-treatment concentrations of these respective biopolymers. This may reflect the complex compositional nature of respiratory aspirates from COVIID-19 ARDS patients and interactions among the various components present. Nonetheless, upon targeting HA and DNA with enzymes we do see a change in modulus which points to an important functional contribution of these polymers to the modulus of these secretions.

Thinning of the fluid to improve lung clearance is a common goal across a range of diseases with respiratory inflammation (57–60). As we observed in our study, treatment of respiratory secretions with an enzyme to digest the biopolymers (and hence decrease the polymer entanglements) will decrease the flow resistance of thick samples with an initial high modulus. The impact of the DNase is established clinically, as it has been used in treating CF lung disease (61, 62) and is under investigation as a treatment for COVID (NCT04359654, NCT04541979); however, the use of HAdase for improving the flow properties of respiratory secretions is a relatively new approach and requires further investigation. More research is needed to identify ideal treatment conditions such as dosages and dosage regimens. Further, targeting the production of the HA during infection may be more successful than relying on a post-production degradation approach. Treatment with a pharmaceutical HA-inhibitor such as hymecromone (4-methylumbiliferone) (63) may be a viable approach to limit the deposition of HA during infection.

We also observed upregulation of many pro-inflammatory cytokines and chemokines in the COVID-19 cohort compared to healthy controls, including IL6, TNF, IL1b, IL18, MCSF, CCL5, CCL4, MIP-1 α /CCL3, and others. These data are consistent with an excessive proinflammatory macrophage activation phenotype and the contribution of myeloid cells to pathogenic inflammation as described by other reports (64–68). This is also consistent with published data correlating abundance of low molecular HA to a hyperinflammatory state (42, 43, 69).

Conversely, we observed significant decrease in IFN-α2 in COVID-19 respiratory secretion samples, compared to the healthy controls. Smith et al. likewise reported significantly lower expression of IFN-α2 in the nasopharynx of patients with COVID-19, compared to healthy controls (70). Hadjadj et al. reported that plasma levels of IFN-α2 decreased with disease severity, but were not significantly different compared to healthy controls (71). Our results are different from a plasma study which reported higher IL33 in severe COVID-19 cohort but our findings are consistent with a nasopharynx study (70, 72). IL-13 was also not significantly different from healthy controls in our samples, in contrast to a previous report (56). These differences may reflect distinct systemic versus local pulmonary immune responses in COVID-19 infection. To our knowledge, this is the first study to measure cytokines, chemokines, and other factors in respiratory secretions from intubated COVID-19 patients.

These studies have several limitations. Most notable is the small numbers of cases and samples of secretions involved. These findings need to be confirmed in larger, multi-center studies involving individuals with diverse backgrounds and case presentations. The underlying mechanisms that lead to increased HA would also benefit from further research to identify the causative cell types and signalling pathways. In addition, data in SARS-CoV-2 animal models would enable improved understanding of the contribution of HA to pathogenesis in this disease. Finally, to safely acquire the rheology data, the COVID-19 samples were heat treated to render the samples non-infectious. In control CF samples, this same heat treatment was found to decrease the modulus (Supplemental Figure 4A), presumably due to the denaturation of biopolymers in the sample. Since we observed that higher modulus samples had larger responses to enzymatic treatment, the true effect of enzymatic treatment on COVID-19 lung secretion may be larger than that reported here using heat-treated samples. Future studies should further evaluate a range of enzymatic treatment dosages and durations to assess the rheological effects on non-heat-treated COVID-19 lung secretions. Additionally, the necessity of using induction to collect healthy sputum is a limitation.

These studies may inform the development of much needed therapeutics for patients with COVID-19. Indeed, a study of oral hymecromone as a potential tool for HA inhibition was recently completed in healthy subjects (NCT02780752); the results of this trial are currently in press (73). Developing treatments that render the respiratory secretions of lungs less viscous, and thus easier to clear via natural mucociliary clearance, could be pivotal to improving clinical outcomes in severe COVID-19 and ARDS.

## Methods

### Histologic staining of lung tissues for HA, versican, and TSG6

For Figure 2 describing HA histologic staining, human lung tissue was obtained from a de-identified autopsy specimen provided through the Stanford Pathology Department in the form of formalin-fixed, paraffin-embedded histologic specimen. Histological staining for HA was performed as described previously (74). In brief, 5-µm thick sections were cut on a Leica RM 2255 Microtome (Leica Microsystems Inc.). For HA affinity histochemistry (AFC) the Bond Intense R Detection kit, a streptavidin-horse radish peroxidase (HRP) system, (Leica Microsystems, Inc.) was used with 4 µg/mL biotinylated-HABP in 0.1 % bovine serum albumin (BSA) in phosphate buffered saline (PBS) as the primary. All images were collected using the BZ-X710 inverted fluorescence microscope (Keyence, Osaka, Japan) at 20X magnification. Montages were generated using the Keyence BZX Analyzer software’s stitching function. For figure 3, HA histological staining was performed similarly, except for the use of 4 PLUS Streptavidin HRP label (Biocare Medical), instead of the Bond Intense R Detection kit.

For figure 3 and supplemental figures, most of the lung tissue samples from healthy, COVID-19 ARDS, and non-COVID-19 ARDS groups were obtained in collaboration with clinical partners at University of Texas Health Science Center at Houston (HSC-MS-15-1049 and HSC-MS-08-0354) and Houston Methodist Hospital (Pro00003392). Discarded donor lungs for transplantation served as healthy controls were obtained from LifeGift Organ Procurement (Houston, TX). Lung tissues were collected from the mid portion of the upper and lower lobes as described previously (75). The details of the study population are summarized in Supplemental Table 1. The CF tissue samples were provided through the Stanford Pathology Department. All histologic specimens were formalin-fixed and paraffin-embedded 5-µm thick sections. HA staining was performed as detailed above. Histological staining for versican and TSG-6 was performed as described previously (76). Rabbit anti-versican antibody, clone EPR12277 (ab177480; Abcam) and rabbit anti-TSG-6 antibody (ab204049; Abcam) were used at 1:50 dilution. Positive and negative controls were included in each staining experiment.

Whole-section imaging was performed using a Aperio (Leica) AT2 Digital Pathology whole slide scanner at the Stanford University Department of Pathology, Human Pathology/Histology Service Center. Slides were scanned in bright-field at a 20× objective and the digital images imported for analysis using the Aperio Imagescope v12.4.3.5008 viewing software. All the images were taken under the same experimental settings. Tissues were also examined using an Amscope T720Q microscope, and higher magnification images (40X) were acquired using Amscope digital camera (MU1403) and imaging software (for Figure 3).

### Histology quantification

150 500 pixel x 500 pixel regions of interest (ROIs) were randomly sampled from each tissue section imaged with the Aperio (Leica) AT2 Digital Pathology whole slide scanner. The RGB image was deconvolved into its hematoxylin and DAB components using the algorithm from (77), implemented in the rgb2hed function in the scikit-image Python package. The frequency of pixels in the DAB channel above a pre-defined threshold was computed per ROI. The threshold was calibrated on tiles randomized across groups to optimally detect positive DAB staining. The mean DAB+ frequency across ROIs was calculated per sample and plotted as % DAB+ area.

### Collection of human respiratory secretions

We collected respiratory secretions from patients enrolled in the Stanford University sputum biobank study from March 2020–March 2021 (IRBs 28205 and 55650). COVID-19 samples were respiratory secretions obtained during the course of routine clinical care. All COVID-19 samples were collected from ventilated patients who were diagnosed with ARDS. Eligibility criteria included admission to Stanford Hospital with a positive SARS-CoV-2 nasopharyngeal swab by RT-PCR. Patients admitted to the ICU were included. Patients were phenotyped for ARDS using the Berlin criteria (acute onset of hypoxemic respiratory failure with a PaO2/FIO2 ratio (i.e., the ratio of the partial pressure of arterial oxygen to the percentage of inspired oxygen) of <300 on at least 5 cm of positive end-expiratory pressure, bilateral infiltrates on chest X-ray). For controls, sputum was collected from asymptomatic adult donors. Healthy control subjects were asymptomatic, aged 24–50 years. Sputum samples from CF patients were collected during routine care. All samples were frozen at -80°C immediately after collection. Samples were thawed slowly on ice then heat treated at 65°C for 30 minutes to render the virus inactive and the sample noninfectious prior to further analyses. These studies were approved under APB protocol #2379.

### Compositional characterization of respiratory secretions

The solids content of the human respiratory secretions was determined by taking the ratio of the freeze-dried mass and wet mass of the samples following at least 2 days of lyophilization. Protein concentrations were determined using the Pierce Bicinchoninic Acid (BCA) Protein Assay (Thermo Scientific) following manufacturer’s instructions. HA concentration was determined using a modified HA Enzyme-Linked Immunosorbent Assay (ELISA) as previously described (77). DNA concentrations were determined using Quant-iT dsDNA Broad-Range Assay Kit (Molecular Probes-Life Technologies) following manufacturer’s instructions. The pH of the samples was measured using pH-indicator strips (Supelco).

### Gel electrophoresis to characterize HA molecular weight

Respiratory secretion samples were treated with 250 U benzonase for 30 min at 37°C for nucleic acid digestion, followed by an incubation with 1 mg/ml proteinase K for 4 hrs at 65°C for further digestion. Proteinase K was heat inactivated by incubating the samples at 100°C for 5 min. Insoluble material was removed by centrifugation at 17,000 g for 10 min before further processing. Samples were precipitated with ethanol overnight at -20°C by adding 4 volumes of pre-chilled 200-proof ethanol to each sample. The following day, the samples were centrifuged at 17,000 g for 10 min. The supernatant was discarded, and the pellet was washed by adding 4 volumes of pre-chilled 75% ethanol. Samples were centrifuged at 17,000 g for 10 min and the resulting pellet air dried at room temperature for 20 minutes. Each sample was resuspended in 100 μl of 100 mM ammonium acetate in water, lyophilized and resuspended in 10 μl of 10 M formamide. Samples were separated on a 1% Tris-acetate EDTA (TAE) agarose gel run at 100 V, then stained with Stains-All (1.25 mg/200 mL in 30% ethanol) (Sigma). The gel was imaged on a BioRad GS-800 Calibrated Densitometer. Twice the volume of healthy control samples was loaded in each gel lane compared to CF and COVID-19 samples.

### Gel electrophoresis to characterize DNA molecular weight

Respiratory secretion samples were mixed with loading solution and separated on a 1% agarose gel with 0.5 µg/mL ethidium bromide at 120 V. Samples were separated on the same gel with a 2-Log DNA Ladder (New England Biolabs). Gels were imaged on a BioRad ChemiDoc MP imaging system.

### Microrheology measurements

Dynamic light scattering (DLS) microrheology data was collected as previously described (46) with minor modifications as described below. Due to the presence of naturally occurring particulates within all samples, no additional beads were required to induce light scattering. Light scattering was collected from a Malvern Nano Zetasizer Nano ZS with a 633 nm laser operated in 173° backscatter mode. The raw intensity autocorrelation function of a respiratory secretion sample was measured at a specified measurement position for 30 minutes at 37°C. Following initial microrheology measurements, the same respiratory secretion sample was then treated with either 1) benzonase nuclease (250 U/mL) for 1 hour at 37°C, 2) hyaluronidase (50 mg/mL, Sigma Aldrich) for 2 hour at 37°C, or 3) 1x phosphate buffered saline for 1 hour at 37°C as a dilution control. After the allotted reaction time, the DLS measured the raw intensity autocorrelation function of the sample at the same settings as before. To safely determine the effect of heat on the rheological behavior of respiratory secretions, we measured the intensity autocorrelation function of CF sputum, which is similar to COVID-19 respiratory secretions in both composition and rheological behavior, before and after the same heat treatment that all COVID-19 respiratory secretion samples were subjected to prior to handling. The heat treatment significantly decreased the resistance to flow (i.e. the elastic modulus) of the CF sputum (Supplemental Figure 4A).

### Microrheology data analysis

The intensity autocorrelation data acquired above was analyzed using the custom analysis package found at dlsur.readthedocs.io. The size of the particulates was assumed to be 500 nm in diameter for all samples. While this assumption affects the absolute value of the modulus derived from the scattering autocorrelation function, it has the same proportional effect across all samples. Thus, the trends observed in the microrheology data, along with the conclusions drawn from those trends, are unaffected by this assumption. All rheological measurements in this study obtained the complex modulus over a wide range of frequencies (from about 10^1^ to 10^6^ Hz), but only the modulus value at one frequency was used when comparing the modulus across samples and conditions. This is a common approach when comparing rheological results of lung secretions (25). To determine this frequency, the complex moduli of the pre- and post-treatment were compared. In the spectrum with the higher complex modulus (typically the pre-treatment), a single frequency was determined by selecting either a) the middle of the “plateau” region of the elastic modulus (Supplemental Figure 3A) or, in the case of no plateau region, b) the lowest frequency for which there is data (Supplemental Figure 3B). Some samples had limiting frequency ranges due to the fast decay of the measured autocorrelation function, which often corresponds to solutions with less resistance to flow. For a single sample, the same frequency was chosen for the pre-treatment modulus and post-treatment modulus. The change in modulus with dilution, ΔG_Saline_, was determined by subtracting the modulus of the sample after dilution to before dilution. The change in modulus with enzyme (DNase or hyaluronidase) treatment, ΔG_enzyme_, was determined by subtracting the modulus of the sample after enzyme addition from the modulus before enzyme addition. The measured moduli pre-treatment, G_pre_, and post-treatment, G_post_, for all individuals are shown in Supplemental Figure 4B.

### Luminex-EMD Millipore Human 80 Plex assays

Luminex assays were performed by the Human Immune Monitoring Center (HIMC) at Stanford University. The COVID-19 respiratory secretion samples and the respective healthy controls were inactivated with 1% Triton-X (vol/vol) (79, 80). Samples were spun down rigorously and diluted 1:6 for the Luminex assay. Kits were purchased from EMD Millipore Corporation and used according to the manufacturer’s recommendations with modifications described as follows: H80 kits include 3 panels: Panel 1 is Milliplex HCYTA-60K-PX48. Panel 2 is Milliplex HCP2MAG-62K-PX23. Panel 3 includes the Milliplex HSP1MAG-63K-06 and HADCYMAG-61K-03 (Resistin, Leptin and HGF) to generate a 9 plex. The assay setup was performed as recommended by the manufacturer. Briefly, samples were mixed with antibody-linked magnetic beads on a 96-well plate and incubated overnight at 4°C with shaking. Incubation steps were performed on an orbital shaker at 500-600 rpm. Plates were washed twice with wash buffer in a Biotek EL×405 washer (BioTek Instruments, Winooski, VT). Following a 1-hour incubation at room temperature with biotinylated detection antibody, streptavidin-PE was added for 30 minutes with shaking. Plates were washed as described above and PBS added to wells for reading in the Luminex FlexMap3D Instrument with a lower bound of 50 beads per sample per cytokine. Custom Assay Chex control beads were purchased and added to all wells (Radix BioSolutions, Georgetown, Texas). Wells with a bead count <50 were flagged, and data with a bead count <20 were excluded. Mean fluorescence intensity (MFI), an estimate of analyte concentration, was used to compare expression in each sample. The cytokines/chemokines/adhesion molecules measured by the Luminex assay are presented in Figure 6 and the raw data are available in the supplemental materials.

### Statistics

Data are expressed as mean +/- SD of n independent measurements. Significance of the difference between the means of two or three groups of data was evaluated using a one-way ANOVA followed by Tukey’s or Dunnett’s post hoc test, as indicated. Student t test with Welch’s correction or Mann Whitney test were used to determine the significance between the means of two groups of data, as indicated. A p value less than <0.05 was considered statistically significant. The small number of samples and large amount of scatter in our rheological data necessitates a statistical measure that can capture the significance of any correlation. Therefore, we chose to use the Bayesian Information Criterion (BIC) as the statistical metric because it can establish correlation while accounting for measurement error in the analysis, which is generally not the case for the more commonly seen R^2^ metric in linear regressions and paired t-tests. More simply put, the BIC tests the significance of any trend in the data relative to there being no trend, and a BIC value greater than 10 shows that the trend is statistically significant relative to the hypothesis that there is no trend in the data.

### Study Approval

All secretion samples were obtained under the auspices of research protocols approved the Stanford Institutional Review Board (IRB) (Stanford IRB approval #28205, #53685, #55650, #37232, and #43805). Samples were collected after written informed consent from patients or their surrogates prior to inclusion in the study.

## Data Availability

All data produced in the present study are available upon reasonable request to the authors.

## Author contributions

G.K., P.L.B., A.J.S, and S.C.H. conceived the study. E.B.B, M.R.N., M.G.O., D.P.R., A.E. P.-N., S.Y., H.K.-Q., A.M.P., B.Z., M.L.B., Stanford COVID Biobank, C.E.M., and A.J.R. identified, enrolled, and consented eligible patients and patient samples. M.J.K., G.K., S.D., and P.L.B. processed patient samples. M.J.K., G.K., S.D., P.C.C., G.L.B., P.Y.J., performed experiments. V.A.d.J.-P. provided the histopathology descriptions and analysis. M.J.K., G.K., S.D., P.C.C., E.B.B., M.P., G.L.B., M.R.N, N.N., R.B.V., T.N.W., C.E.M., A.J.R., A.J.S., S.C.H., and P.L.B. performed data analysis and interpreted data. M.J.K., G.K., S.D., C.E.M., A.J.S., S.C.H., and P.L.B. wrote the manuscript with input from all authors. Authorship order for co-first authors was determined via mutual agreement between M.J.K., G.K., and S.D.

## Acknowledgements

We are grateful to all participants in this study. Our thanks go to A. Wardle for his reading of the manuscript and his helpful comments. We would also like to thank G. Nolan for providing access to the Keyence microscope and analysis software. Thank you to the Human Immune Monitoring Center (HIMC) at Stanford University – ImmunoAssay Team for their help on the Luminex assays. This research program is supported by a grant from the Stanford Innovative Medicines Accelerator program and the COVID-19 Response program from Stanford ChEM-H (Chemistry, Engineering & Medicine for Human Health). The Stanford COVID-19 Biobank Study Group and A.J.R. are funded by NIH/ NHLBI K23 HL125663. A.J.R. was supported by NIH grant T32 AI007502-23. See Supplemental Acknowledgments for Stanford COVID-19 Biobank Study Group details.

