## Supplemental Files for "Biochemical, Biophysical, and Immunological Characterization of Respiratory Secretions in Severe SARS-CoV-2 (COVID-19) Infections"

### Table of Contents

Supplemental Table 1. Demographic details of healthy cohort

Supplemental Figure 1. HA molecular weight sizing gel

Supplemental Figure 2. HA accumulation and distribution in COVID-19 ARDS, non-COVID ARDS, CF and healthy lung sections.

Supplemental Figure 3. Versican accumulation and distribution in COVID-19 ARDS, non-COVID ARDS, CF and healthy lung sections.

Supplemental Figure 4. TSG6 accumulation and distribution in COVID-19 ARDS, non-COVID ARDS, CF and healthy lung sections.

Supplemental Figure 5. DNA molecular weight sizing gel

Supplemental Figure 6. Pre-treatment and post-treatment modulus identification

Supplemental Figure 7. Microrheology data analysis

Supplemental Figure 8. Impact of concentration of DNA and HA on the modulus change due to enzymatic treatment

Supplemental Figure 9. MUC5AC and MUC5B Western blot analysis of COVID-19 respiratory secretions

Supplemental Figure 10. Heat map of raw mean fluorescence intensity (MFI) data scaled per row

Supplemental Figure 11. Immunological characterization of respiratory secretions from CF patients.

Supplemental Methods. Western blot analysis to characterize mucins

Supplemental Acknowledgments

Supplemental Table 1. Demographic details of healthy cohort

| **Characteristic** | **Adult cohort** *(n=6)* |
|---|---|
| Age (years) | 42.8 (31 - 58) |
| Male gender | 5 (83.3%) |
| Race/Ethnicity | Non-black Hispanic 1 (16.7%)<br>White 5 (83.3%) |
| Preexisting diabetes mellitus | 0 (0%) |
| Preexisting pulmonary disease | 0 (0%) |
| Preexisting cardiac disease | 0 (0%) |

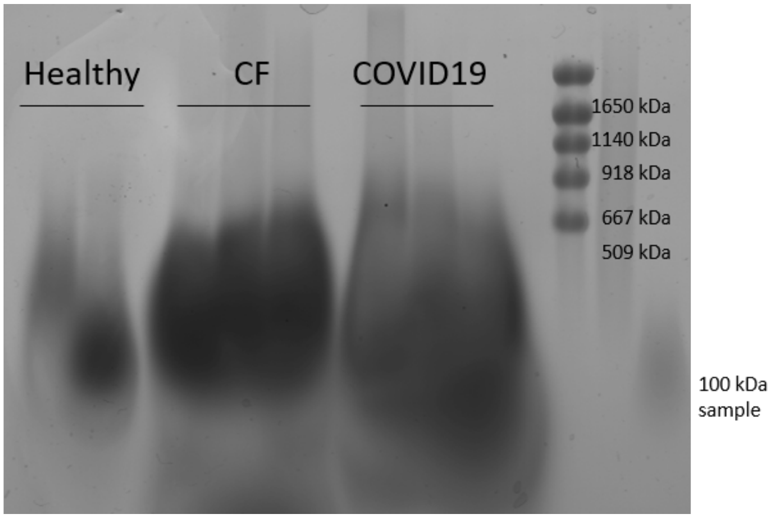


**Supplemental Figure 1.** Representative HA molecular weight sizing gel from Healthy, CF, and COVID-19 respiratory secretion samples.

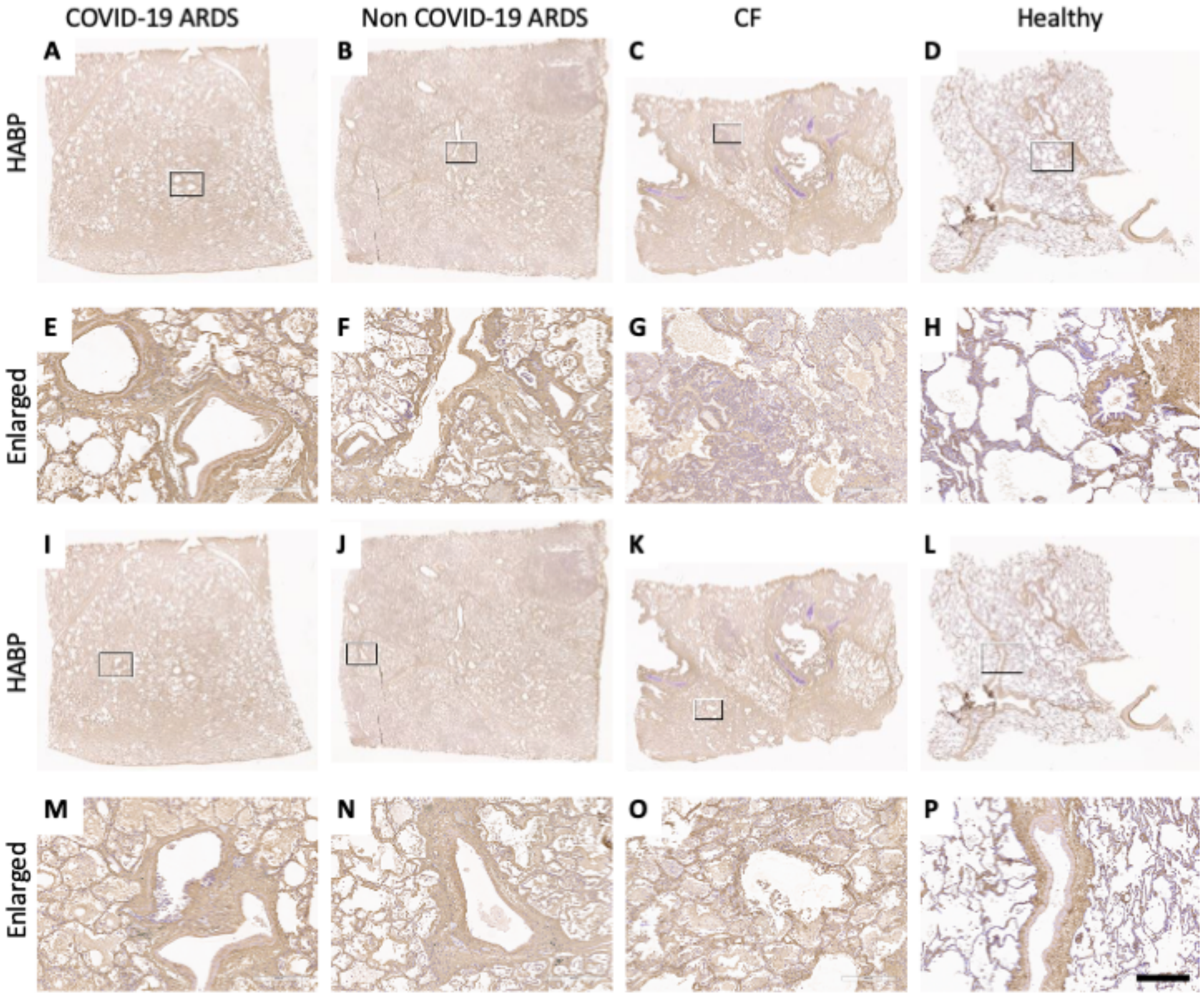


**Supplemental Figure 3. HA accumulation and distribution in COVID-19 ARDS, non-COVID ARDS, CF and healthy lung sections.** Representative histological cadaveric lung sections from donors with COVID-19 ARDS, donors with non-COVID-19 ARDS, donors with CF, and healthy donors stained with (**A-P**) HABP. (**E-H**) and (**M-P**) are enlarged sections from (**A-D**) and (**I-L**) respectively (box highlights enlarged area). Nuclei are stained in blue, and HABP is stained in brown. Scale bar A-P 400 µm. Whole-section imaging was performed using a Aperio (Leica) AT2 Digital Pathology whole slide scanner. Slides were scanned in bright-field at a 20× objective and the digital images imported for analysis using the Aperio Imagescope v12.4.3.5008 viewing software.

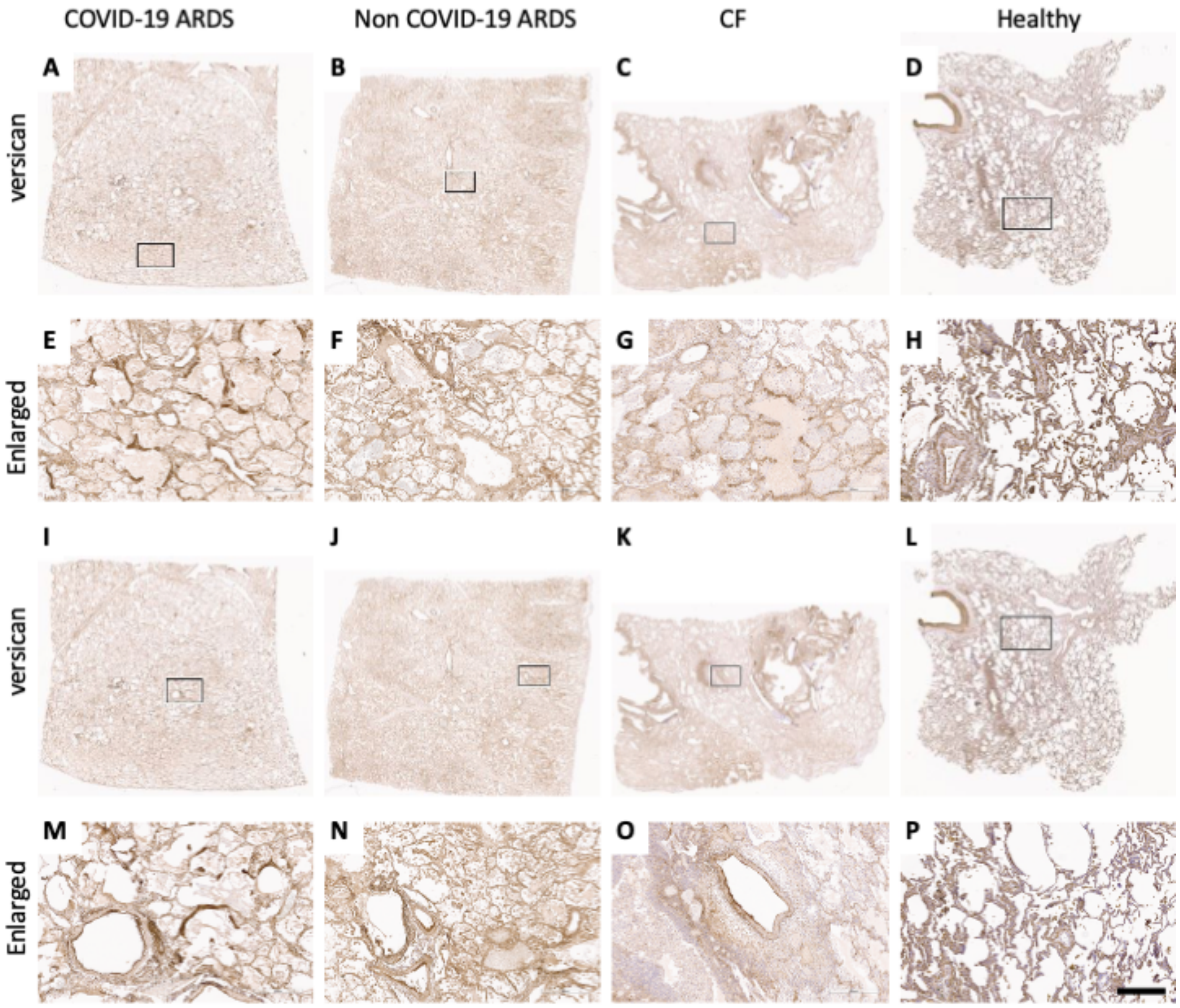


**Supplemental Figure 4. Versican accumulation and distribution in COVID-19 ARDS, non-COVID ARDS, CF and healthy lung sections.** Representative histological cadaveric lung sections from donors with COVID-19 ARDS, donors with non-COVID-19 ARDS, donors with CF, and healthy donors stained with (**A-P**) Versican. (**E-H**) and (**M-P**) are enlarged sections from (**A-D**) and (**I-L**) respectively (box highlights enlarged area). Nuclei are stained in blue, and HABP is stained in brown. Scale bar A-P 400 μm. Whole-section imaging was performed using a Aperio (Leica) AT2 Digital Pathology whole slide scanner. Slides were scanned in bright-field at a 20× objective and the digital images imported for analysis using the Aperio Imagescope v12.4.3.5008 viewing software.

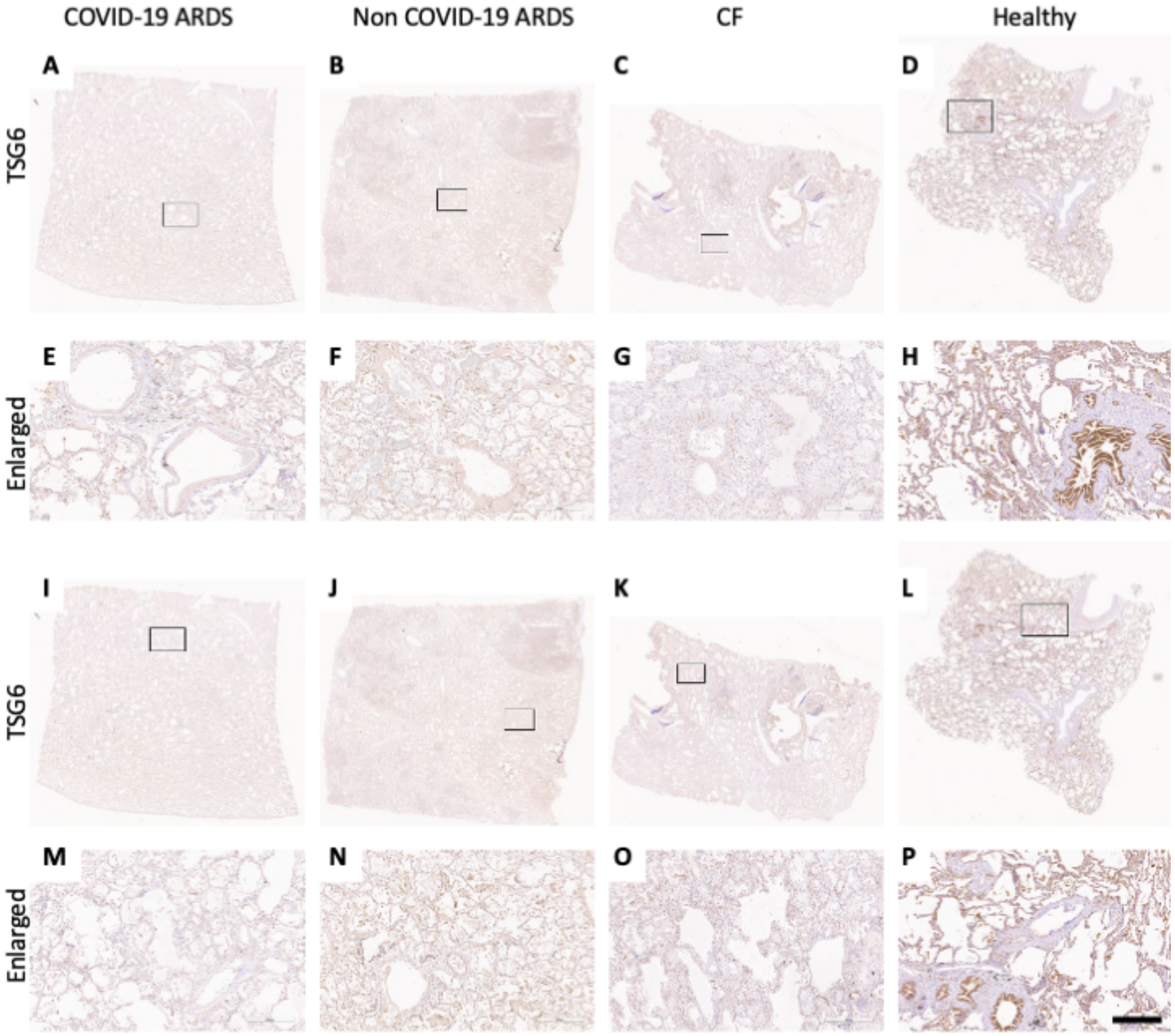


**Supplemental Figure 5. TSG6 accumulation and distribution in COVID-19 ARDS, non-COVID ARDS, CF and healthy lung sections.** Representative histological cadaveric lung sections from donors with COVID-19 ARDS, donors with non-COVID-19 ARDS, donors with CF, and healthy donors stained with (**A-P**) TSG6. (**E-H**) and (**M-P**) are enlarged sections from (**A-D**) and (**I-L**) respectively (box highlights enlarged area). Nuclei are stained in blue, and HABP is stained in brown. Scale bar A-P 400 µm. Whole-section imaging was performed using a Aperio (Leica) AT2 Digital Pathology whole slide scanner. Slides were scanned in bright-field at a 20× objective and the digital images imported for analysis using the Aperio Imagescope v12.4.3.5008 viewing software.

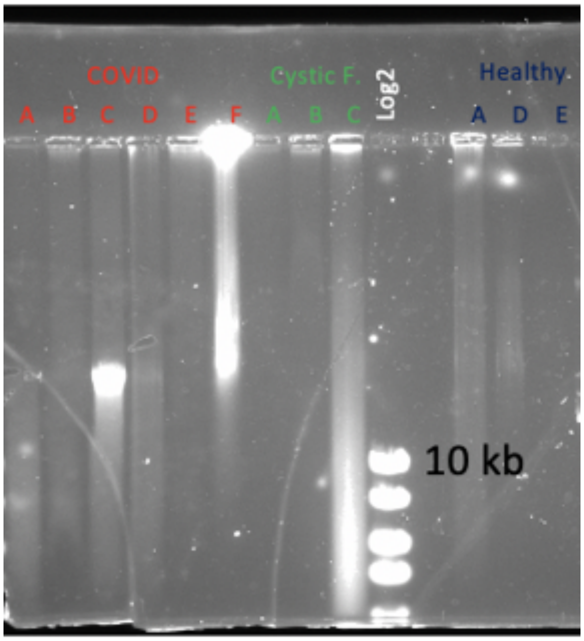


**Supplemental Figure 5.** Representative DNA molecular weight sizing gel from Healthy, CF, and COVID-19 respiratory secretion samples.

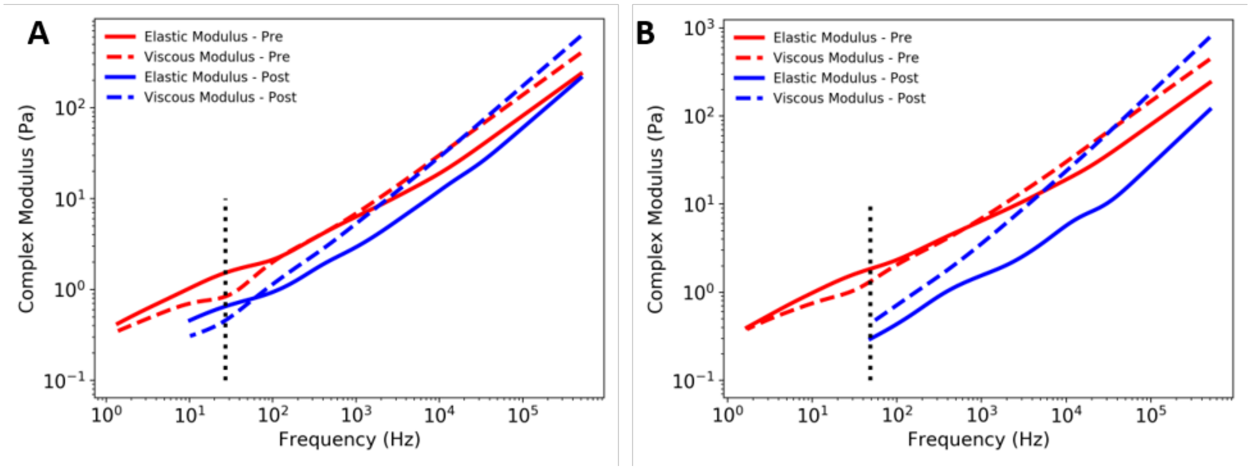


**Supplemental Figure 6.** Representative rheological spectra for pre-treatment and post-treatment samples, where the dotted line represents the frequency chosen at which the single modulus value was used for comparison when (**A**) a "plateau" region of the elastic modulus exists or, (**B**) in the case of no plateau region and hence the lowest frequency is selected.

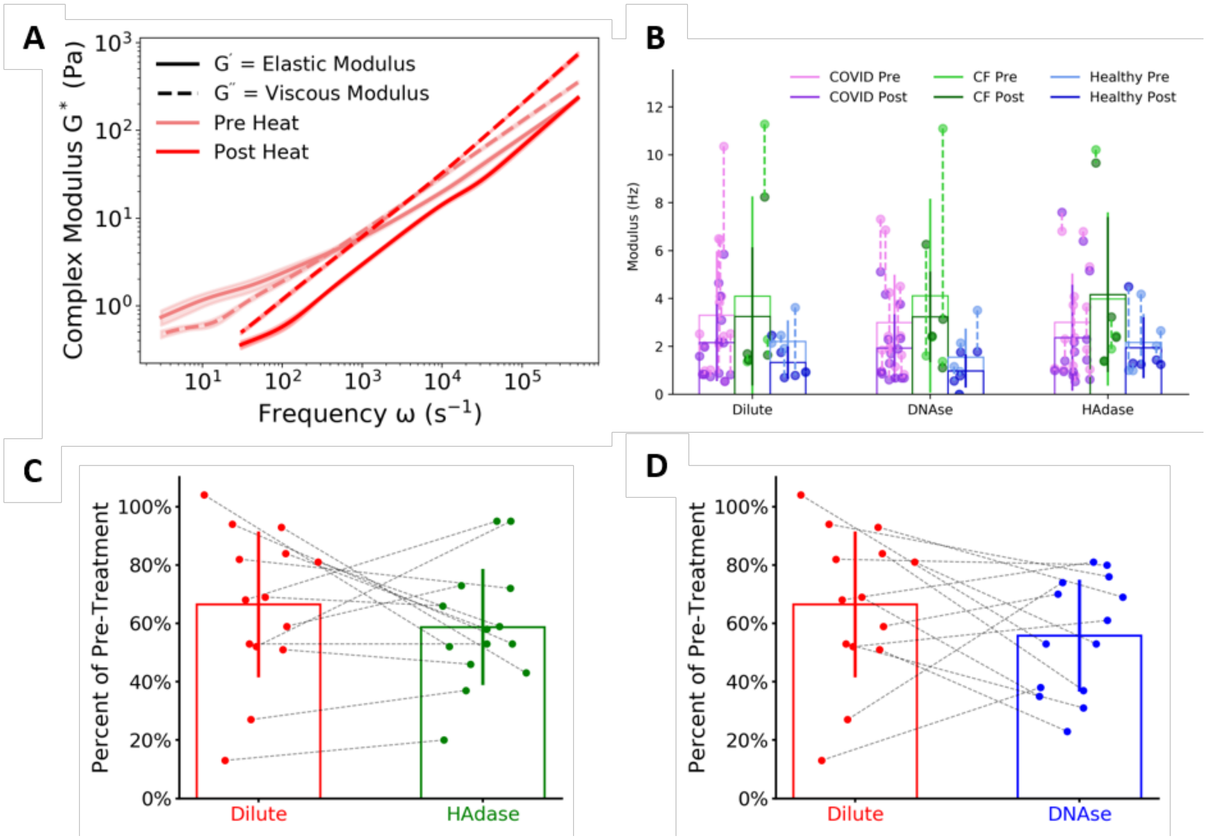


**Supplemental Figure 7.** (**A**) Microrheology data showing the impact of virus-killing heat treatment on cystic fibrosis (CF) respiratory secretions. (**B**) A plot of the measured moduli of all samples tested. Lighter and darker data points indicate the pre- and post-treatment, respectively, with a dashed line connecting data from the same patient sample. (**C**) A plot of relative change in modulus of COVID-19 respiratory secretions after dilution or HAdase enzymatic treatment compared to pre-treatment modulus, with a dashed line connecting data points generated from the same patient sample (n=14). (**D**) A plot of relative change in modulus of COVID-19 respiratory secretions after dilution or DNase enzymatic treatment compared to pre-treatment modulus, with a dashed line connecting data points generated from the same patient sample (n=14).

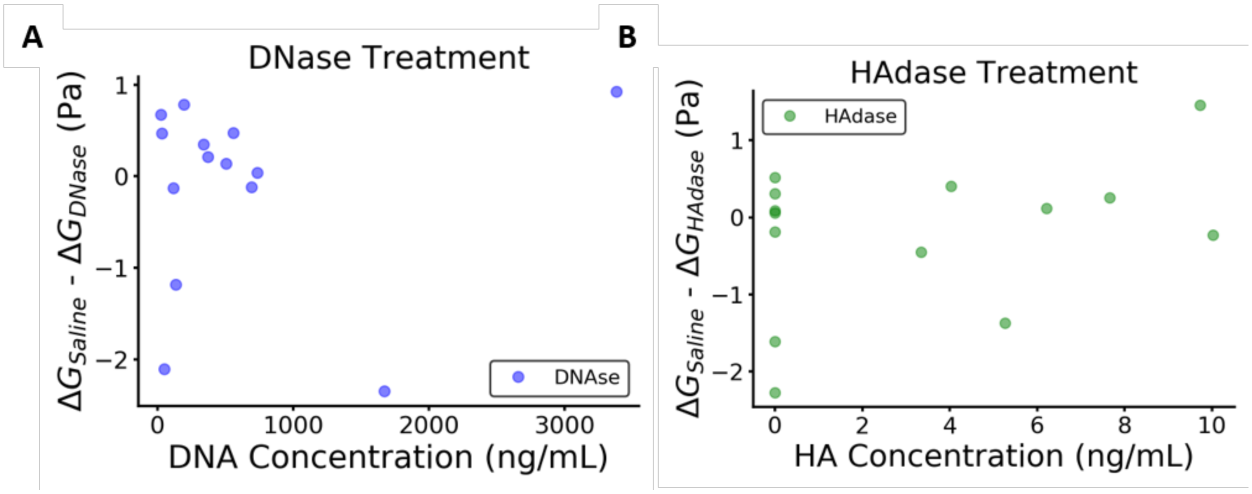


**Supplemental Figure 8.** Impact of concentration of DNA and HA on the modulus change due to enzymatic treatment**.** (**A**) The difference in modulus of COVID-19 lung secretions upon control saline dilution and enzymatic DNAase treatment ($\Delta G_{Saline}$-$\Delta G_{DNase}$) versus the initial, pre-treatment modulus (blue) (n=14). (**B**) The difference in modulus of COVID-19 lung secretions upon control saline dilution and enzymatic HAdase treatment ($\Delta G_{Saline}$-$\Delta G_{HAdase}$) versus the initial, pre-treatment modulus (green) (n=14). We found no strong correlation between biopolymer concentration and modulus change as indicated by the Bayesian Information Criterion (BIC) values being less than 10 (3.5 and 8.5 for DNA and HA).

A

B

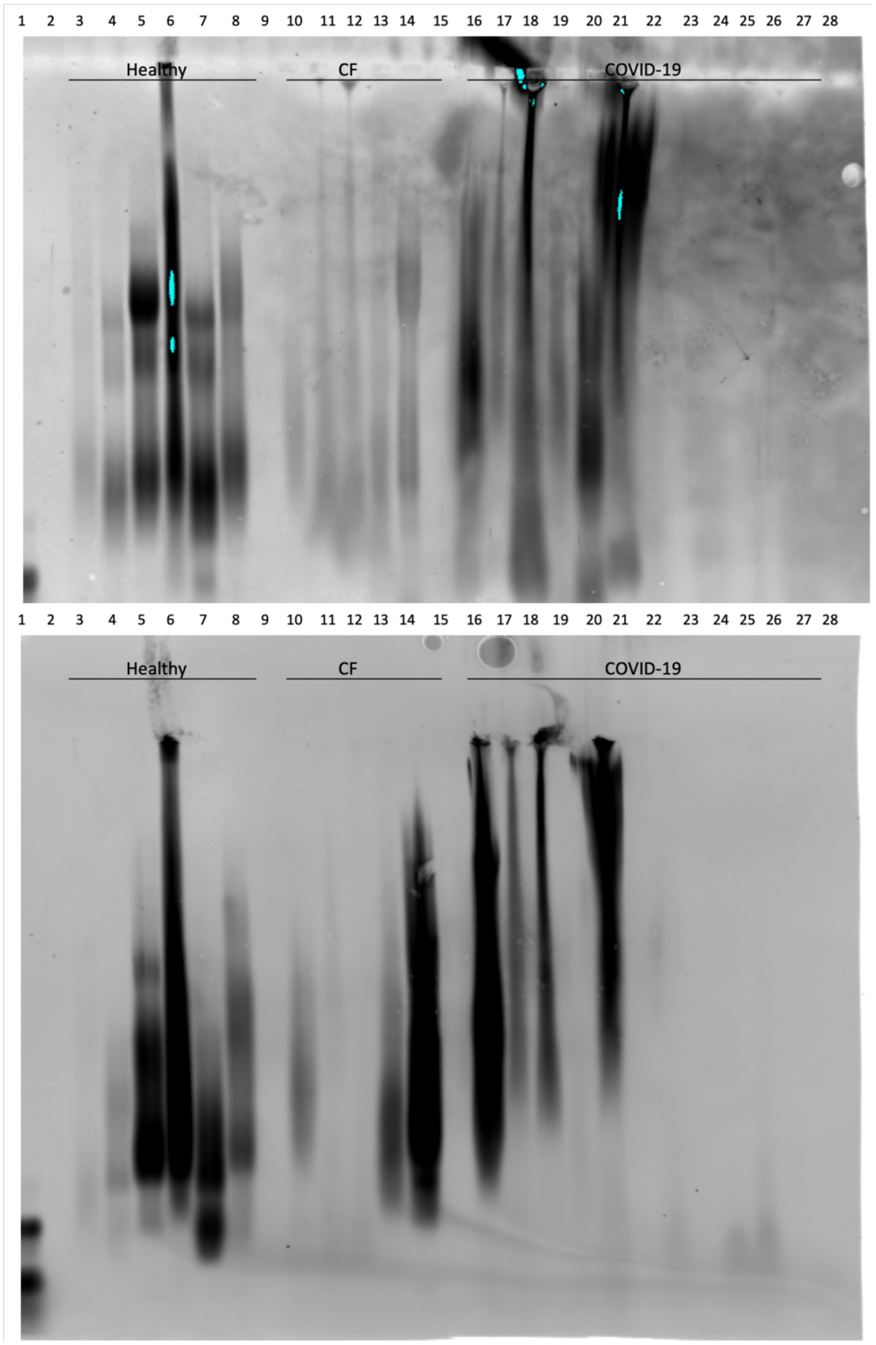


**Supplemental Figure 9.** Representative western blot analysis of MUC5AC and MUC5B protein expression from Healthy (n-6), CF (n=5), and COVID-19 respiratory secretion samples (n=12 for A, n=11 for B). Samples were separated via mucin agarose Western blotting and the membrane was probed with a (**A**) rabbit MUC5AC polyclonal antibody (**B**) mouse MUC5B (A-3) monoclonal antibody. In panel A, lanes 2, 9, 15, 22, 28 are blank, while in panel B lanes 2, 9, 15, 21, 28 are blank.

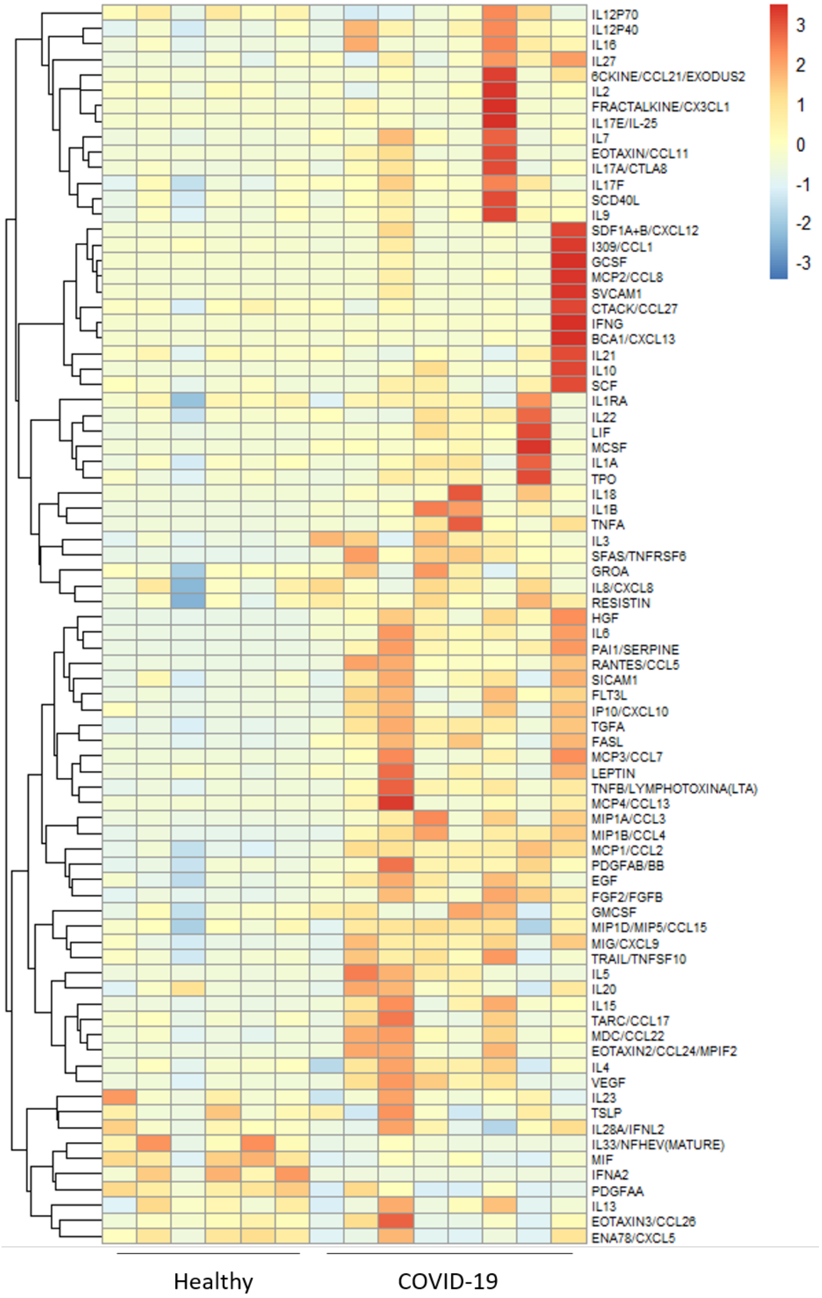


**Supplemental Figure 10.** Heat map of raw mean fluorescence intensity (MFI) data scaled per row (Healthy vs COVID-19). Cytokines, chemokines, adhesion molecules, and growth factors were measured in the respiratory secretion samples of healthy controls (n = 6) and in COVID-19 ARDS patients (n=8) using a bead-based multiplexed immunoassay system, Luminex-EMD Millipore Human 80 Plex assays. Upregulated cytokines are shown in orange and downregulated in blue.

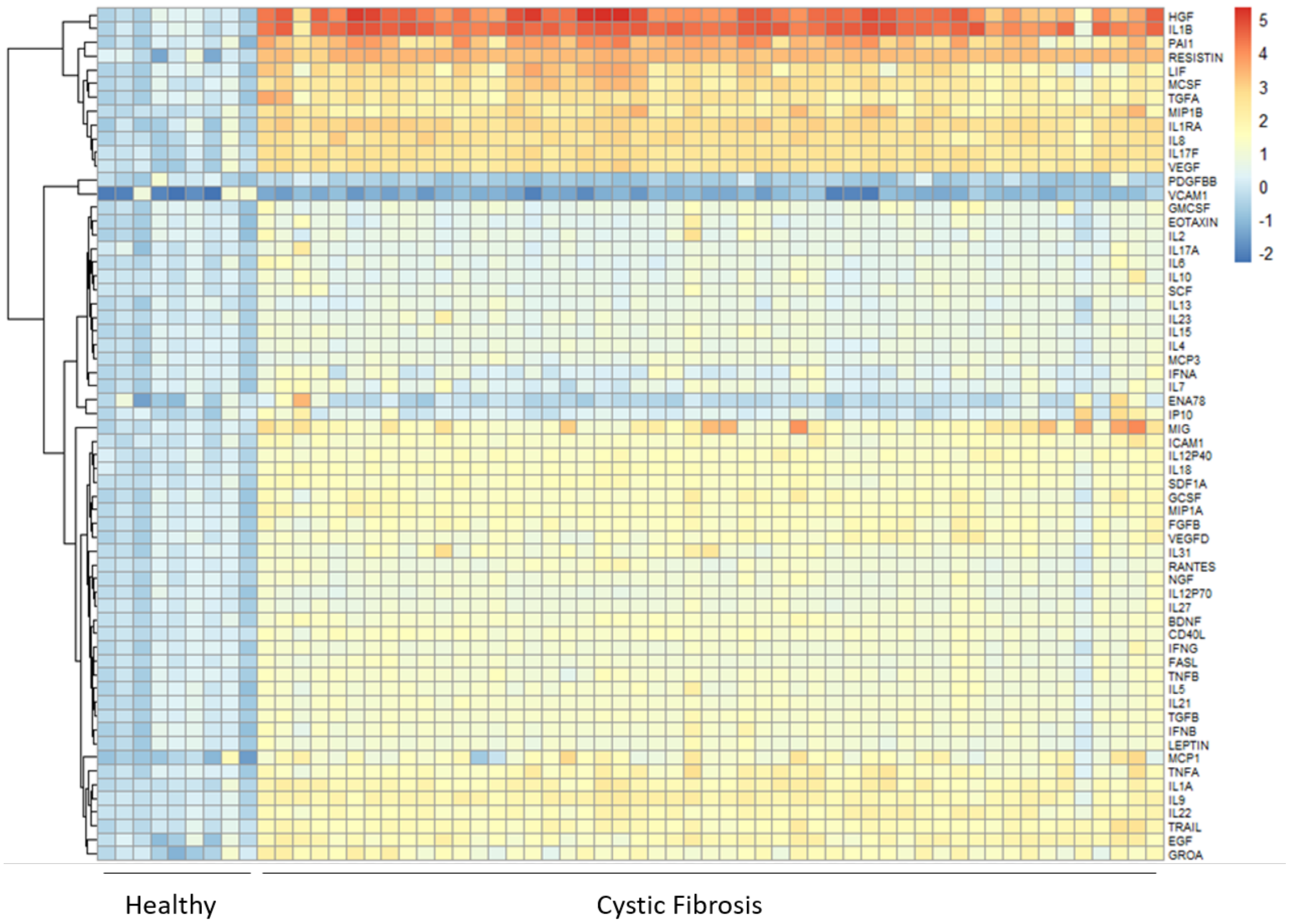


**Supplemental Figure 11.** Immunological characterization of respiratory secretions from CF patients. Heat map of mean fluorescence intensity (MFI) data, log2 transformed, and normalized to average of the healthy controls per cytokine. Data are ordered by KNN clustering of the cytokines (y-axis). Cytokines, chemokines, adhesion molecules, and growth factors were measured in the respiratory secretion samples of healthy controls (n = 9) and in adult CF patients (n=51) using a bead-based multiplexed immunoassay system, Luminex-EMD Millipore Human 63 Plex assays. Upregulated cytokines are shown in orange and downregulated in blue.

### Supplemental Methods

**Western blot analysis to characterize mucins:** Respiratory secretion samples, normalized by weight, were mixed with loading solution, and subjected to Western blot analysis as previously described (doi:10.3791/54153). Briefly, the samples were separated on a 1% agarose gel in 1X TAE-0.1% SDS at 80 V, transferred to nitrocellulose membranes and blocked in 2% Casein, 1% BSA solution. Mucin 5B (MUC5B) was detected with the mouse MUC5B (A-3) monoclonal antibody (sc-393952, Santa Cruz Biotechnology) at 1ug/ml, followed by donkey anti-mouse AF800 secondary antibody (1:10,000). Mucin 5AC (MUC5AC) was detected with the rabbit MUC5AC polyclonal antibody (Thermofisher Scientific) at 0.5ug/ml, followed by goat anti-rabbit AF800 secondary antibody (1:10,000). The blots were imaged on a LI-COR Odyssey Western Blot Imaging system.

**Supplemental Acknowledgements**

The Stanford COVID-19 Biobank Study Group is: Rosen Mann, Anita Visweswaran, Thanmayi Ranganath, Jonasel Roque, Monali Manohar, Hena Naz Din, Komal Kumar, Kathryn Jee, Brigit Noon, Jill Anderson, Bethany Fay, Donald Schreiber, Nancy Zhao, Rosemary Vergara, Catherine Blish, Kari Nadeau, Andra Blomkalns, Ruth O'Hara.
